# Th1 Dominant Nucleocapsid and Spike Antigen-Specific CD4+ and CD8+ Memory T Cell Recall Induced by hAd5 S-Fusion + N-ETSD Infection of Autologous Dendritic Cells from Patients Previously Infected with SARS-CoV-2

**DOI:** 10.1101/2020.11.04.20225417

**Authors:** Peter Sieling, Lise Zakin, Annie Shin, Brett Morimoto, Helty Adisetiyo, Hermes Garban, Philip Liu, Adrian Rice, Justin Taft, Roosheel Patel, Sofija Buta, Marta Martin-Fernandez, Dusan Bogunovic, Elizabeth Gabitzsch, Jeffrey T. Safrit, Lennie Sender, Patricia Spilman, Shahrooz Rabizadeh, Kayvan Niazi, Patrick Soon-Shiong

## Abstract

To address the need for a safe, efficacious vaccine against SARS-CoV-2 infection with the critical properties of enabling both blocking viral entry into cells and clearing virus from cells already infected, we have developed a bivalent, human adenovirus serotype 5 (hAd5) SARS-CoV- 2 S-Fusion + N-ETSD vaccine that is currently in clinical testing. This vaccine uses the next-generation hAd5 [E1-, E2b-, E3-] platform previously used successfully in cancer patients with pre-existing adenovirus immunity, engineered to express both SARS-CoV-2 spike (S) protein modified to improve the generation of neutralizing antibodies to block entry of the virus, and nucleocapsid (N) protein with an Enhanced T cell Stimulation Domain (ETSD) to activate CD4+ and CD8+ T cells to clear the virus and block replication by killing infected cells. The targeting of N to endosomes and lysosomes to enhance CD4+ and CD8+ T-cell responses distinguishes our vaccine. In our previously reported pre-clinical studies we showed that in mice, the hAd5 S-Fusion + N-ETSD vaccine elicits both humoral and T-cell responses that are robust and T helper cell 1 (Th1) dominant. Here we report that the hAd5 S-Fusion + N-ETSD vaccine is recognized by anti-sera and T cells from previously SARS-CoV-2 infected patients, and that the presence of N is vital for T-cell recall. The findings presented herein: (i) demonstrate specific recognition of hAd5 S-Fusion + N-ETSD infected cells by plasma antibodies from previously SARS-CoV-2 infected patients, but not antibodies from virus-naïve subjects; (ii) show enhanced binding of plasma SARS-CoV-2 antibodies from previously infected patients to monocyte-derived dendritic cells (MoDCs) expressing the hAd5 S-Fusion + N-ETSD vaccine as compared to hAd5 S-Fusion alone; (iii) reveal N-ETSD localizes to vesicles associated with MHC class II antigen presentation, including endosomes, lysosomes and autophagosomes in MoDCs; (iv) demonstrate endosome/lysosome-targeted N-ETSD elicits higher interferon-γ T-cell responses than cytoplasm-localized N; and (v) N-ETSD alone or in the hAd5 S-Fusion + N-ETSD construct induces both CD4+ and CD8+ T cell memory recall. This recognition of hAd5 S-Fusion + N-ETSD vaccine antigens by T cells from previously SARS-CoV-2 infected patients, together with the ability of this vaccine candidate to elicit *de novo* immune responses in naïve mice suggests that it recapitulates the natural immune response to SARS-CoV-2 to activate both B and T cells towards viral neutralization and recognition of infected cells, critical for prevention of COVID-19 disease. Intriguingly, our hAd5 S-Fusion + N-ETSD T-cell biased vaccine has the potential to not only provide protection for uninfected individuals, but also to be utilized as a therapeutic for already infected patients to induce rapid clearance of the virus by activating T cells to kill the virus-infected cells, thereby reducing viral replication and lateral transmission.

## INTRODUCTION

The COVID-19 pandemic has resulted in over 1 million fatalities worldwide and has the potential to produce significantly higher numbers of casualties in the near future. The majority of current prophylactic anti-SARS-CoV-2 vaccines under development are designed to prevent further disease-related mortality and morbidity by targeting the viral spike (S) protein with the goal of generating neutralizing antibody responses in recipients prior to viral exposure. However, recent characterization of COVID-19 patient immune responses to SARS-CoV-2 indicates that other immune cells such as T cells are critical to clearing infection and producing long-term immunity to coronavirus infections.^1-3^ Both CD4+ and CD8+ T cells underpin durable humoral responses because CD4+ T cells, while not effector cells like CD8+ T cells, are critical to the generation of robust and long-lasting immunity afforded by antibody-secreting plasma cells and the elimination of infected cells by memory cytotoxic CD8+ T cells. ^4,5^

In response to the dire need for a vaccine to address the global COVID-19 pandemic, ^6-11^ we designed and developed a dual-antigen candidate vaccine with the potential to more broadly activate the immune system to combat SARS-CoV-2, by the inclusion of a modified viral nucleocapsid (N) antigen, a potent CD4+ and CD8+ T cell target, along with an optimized S protein (S-Fusion) to stimulate humoral responses. As previously reported in Rice *et al*., ^12^ to design our COVID-19 vaccine, we leveraged our human adenovirus serotype 5 (hAd5) E1, E2b, and E3 region-deleted [E1-, E2b-, E3-] vaccine platform (Fig. 1A) that is superior to the adenovirus platforms ^13,14^ used in other COVID-19 vaccines currently in clinical trials because it is effective in the presence of pre-existing adenovirus immunity and has a reduced likelihood of generating a vector-targeted host immune response, thus can be used as both the prime and boost. ^15-20^ Using this platform, we constructed a vaccine that comprises the optimized S surface protein, ^21-25^ S-Fusion, to increase cell-surface display and humoral responses; as well as the highly conserved and antigenic N protein found within the viral particle, here with subcellular compartment targeting sequences for enhanced antigen presentation (Fig. 1B). ^26-31^ Overall, we predict this strategy will be safe ^19,32^ and robust in eliciting humoral and T cell responses to SARS-CoV-2.

**Fig. 1.**
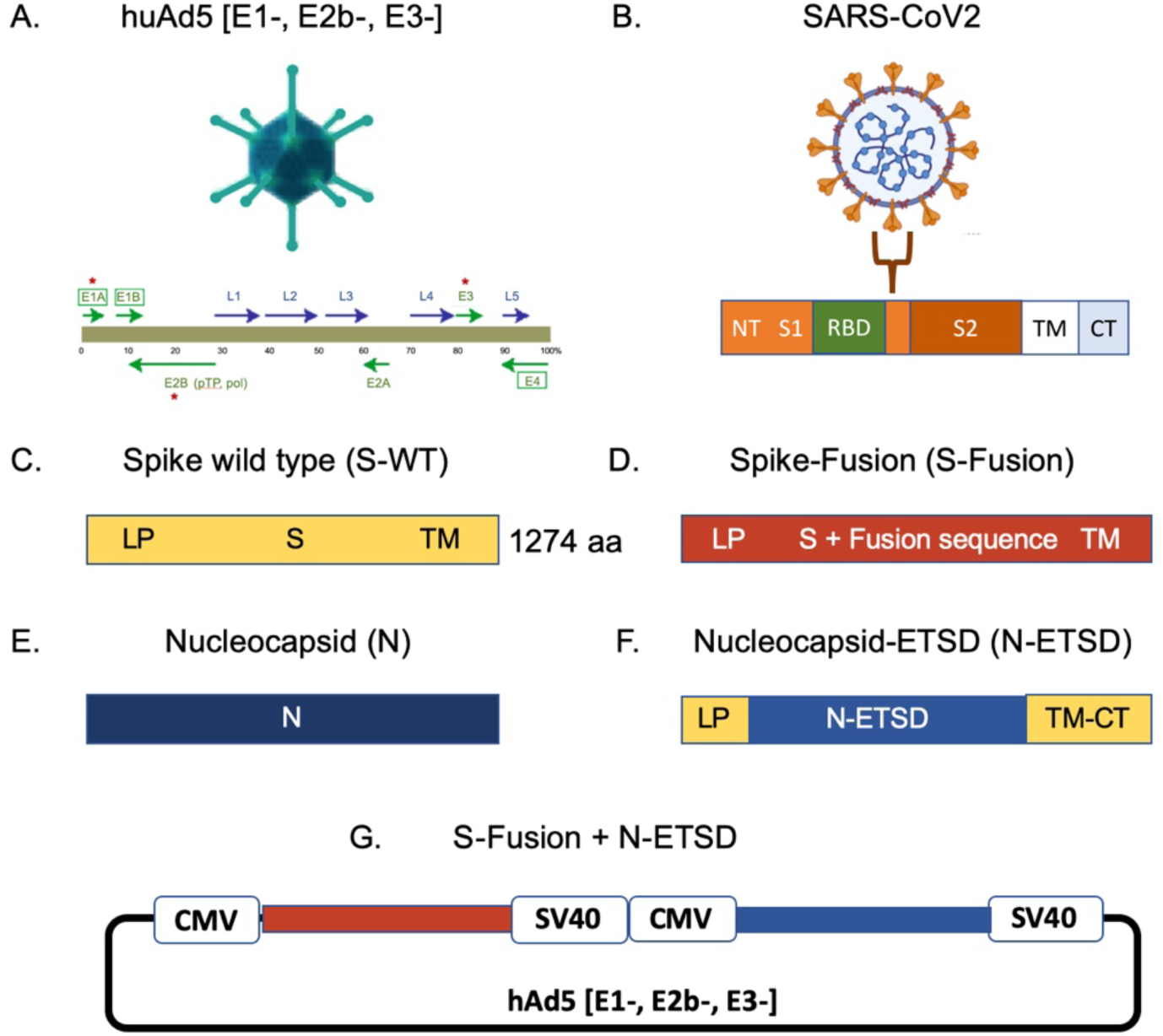
The hAd5 vector, SARS-CoV-2, spike, and constructs. (A) The human adenovirus serotype 5 with E1, E2b, and E3 regions deleted (hAd5 [E1-, E2b-, E3-]) is shown. (B) The SARS-CoV-2 virus displays spike (S) protein as a trimer on the viral surface. S protein comprises the N-terminal (NT), the S1 region including the Receptor Binding Domain (RBD), the S2 and transmembrane (TM) regions, and the C-terminal (CT); other function regions not labeled. (C) Spike wild type (S-WT), (D) spike fusion (S-Fusion), (E) nucleocapsid without ETSD and predominantly cytoplasmic localization (N); (F) N with the Enhanced T-Cell Stimulation Domain (N-ETSD), and (G) the bivalent S-Fusion + N-ETSD constructs are shown.

The addition of N addresses the risk of loss of vaccine efficacy for S-only monovalent vaccines due to the emergence of mutations in S in the population over time (https://nextstrain.org/ncov/global). ^33,34^ In contrast, N is highly conserved ^28^ with a lower risk of mutation, while also being highly immunogenic. ^35-37^ It is a known target antigen for natural immunity, with antibodies and T cells against N being found in the majority of persons recovered from SARS-CoV-2 and similar virus SARS-CoV infections. ^27,35,37-45^ In our vaccine, to optimize the presentation of N, we further added an Enhanced T cell Stimulation Domain (ETSD) to direct N protein to the endosomal-lysosomal subcellular compartment after translation to support MHC class II presentation for T helper cell activation and promotion of CD8+ T cell activation through dendritic cell licensing. ^46^

Despite the risk of emerging mutations, S remains a key antigen for vaccination due to its role in infection. Spike, displayed as a trimer on the viral surface (Fig. 1B), has a Receptor Binding Domain (RBD) that interacts with host angiotensin-converting enzyme 2 (ACE2) to facilitate entry into the host cells and propagation, ^47-49^ thus antibodies against S are key to neutralization of infection. ^50^ Antibodies against S RBD are commonly found in patients recovered from COVID-19 ^51^ as are antibodies against other S epitopes. ^52,53^ In our bivalent vaccine construct, S-Fusion is S optimized by addition of a fusion linker to display the S RBD in a physiologically-relevant form on the cell surface with the goal of improving generation of anti-S RBD antibodies that will be virus neutralizing.

In our design of the bivalent hAd5 S-Fusion + N-ETSD vaccine, we have placed particular importance on its ability to generate T-cell responses. Vaccines currently in clinical trials focus on generating humoral responses as a means to neutralize infection. However, given that antibodies, even if successfully generated and sufficiently neutralizing, may wane over time, ^43,54,55^ the T cell response becomes critical.^2,56-58^ If T cell responses are absent due to virus-induced lymphopenia, even in the presence of abundant neutralizing antibodies, an infected person is at risk for developing acute symptoms of the disease.^59^ While it cannot be excluded that S (and other viral proteins) can induce T-cell responses, the evidence in the literature support a key role for N. Not only have T cell responses to N been found in the majority of patients recovered from COVID-19, ^2^ these responses to N in patients exposed to very similar virus SARS-CoV are remarkably durable. ^2^ Compelling evidence of the importance of N in natural T cell immunity can be found in the recent report of Ferretti *et al*.^60^ who found, using an unbiased genome-wide screen for the precise peptide sequences recognized by memory CD8+ T cells of COVID-19 patients, that only 3 of the 29 shared epitopes were from the spike protein, whereas the highest density of epitopes was located in the nucleocapsid protein. Thus, in the hAd5 S-Fusion + N-ETSD vaccine, N is expected to not only elicit a humoral response, but also a T-cell response that better recapitulates disease-limiting natural immunity.

In our initial report, ^12^ we showed the hAd5 S-Fusion + N-ETSD vaccine provided enhanced cell-surface expression of S RBD that was readily recognized by ACE2, reflecting its conformational integrity; we also demonstrated that N-ETSD with endosomal/lysosomal localization for enhanced antigen presentation generated both neutralizing antibody and CD4+/CD8+ T-cell-mediated responses with Th1 predominance in inoculated mice. Here, we extend those findings using plasma and monocyte-derived dendritic cells (MoDCs) from previously SARS-CoV-2 infected patients to confirm native S antigen expression. We elucidate N-ETSD localization in antigen-presenting MoDCs, showing that it localizes to endosomes, lysosomes, and autophagosomes. Studies have shown that protein processing through this subcellular pathway enhances MHC class II presentation and increases peptide recycling to also enable MHC class I presentation. We demonstrate that by localizing the nucleocapsid protein to the late lysosome-autophagosome compartment both CD4^+^ and CD8^+^ SARS-CoV-2 specific memory T cells are recalled from patients previously infected with SARS-CoV-2. Moreover, in these immune-response recall studies, we show that *in vitro*, hAd5-infected MoDCs presenting S-Fusion and N-ETSD elicit a predominant Th1 response from autologous memory T cells of previously SARS-CoV-2 infected patients. We demonstrate that N in particular drives the CD8+ T cell responses in *in vitro* recall studies. Recapitulation of natural infection and immunity, to the degree it can be achieved by vaccination, by the hAd5 S-Fusion + N-ETSD vaccine makes it a prime candidate for clinical testing of its ability to protect individuals from SARS-CoV-2 infection and COVID-19 and this second-generation vaccine construct has now entered into Phase I clinical trials.

## RESULTS

### The hAd5 [E1-, E2b-, E3-] platform and constructs

For studies here, the next generation hAd5 [E1-, E2b-, E3-] vector was used (Fig. 1A) to create viral vaccine candidate constructs. A variety of constructs were created:

C. S WT: S protein comprising 1274 amino acids and all S domains: extracellular (1-1213), transmembrane (1214-1234), and cytoplasmic (1235-1273) (Unitprot P0DTC2);
D. S-Fusion: S optimized to enhance surface expression and display of RBD;
E. N (N without ETSD): Nucleocapsid (wild type) sequences with tags for immune detection, but without ETSD modification, and predominantly cytoplasmic localization.
F. N with the Enhanced T cell Stimulation Domain (N-ETSD): Nucleocapsid (wild type) with ETSD to direct lysosomal/endosomal localization and tags for immune detection; and
G. The Bivalent hAd5 S-Fusion + N-ETSD vaccine.

### Nucleocapsid antigen engineered with an Enhanced T cell Stimulation Domain (ETSD) directs N to endosomes, lysosomes and autophagosomes in MoDCs, driving enhanced CD4+ T-cell activation

The hAd5 bivalent vaccine construct includes sequences designed to target N to MHC class II antigen loading compartments. To further investigate factors that affect antigen presentation in MoDCs, we evaluated the localization of N-ETSD in human MoDCs compared to its untargeted cytoplasmic counterpart (N). MoDCs from healthy subjects were infected with hAd5 N-ETSD or hAd5 N and localization was determined by immunocytochemistry. N-ETSD showed localization to discrete vesicles, some coincident with CD71, a marker of recycling endosomes (Fig. 2A-C), and LAMP-1, a marker for late endosome/lysosomes (Fig. 2G-I), whereas N was expressed diffusely and uniformly throughout the cytoplasm (Fig. 2D-F; J-L). Studies have shown that lysosomes fuse with autophagosomes to enhance peptide processing and MHC class II presentation. Thus we examined whether N-ETSD localized in autophagosomes in MoDCs with the addition of co-labeling with the autophagosome marker LC3a/b ^61^ to identify another potential site of localization relevant to MHC class II antigen presentation. ^62^ We found that N-ETSD also displayed some co-localization with the autophagosome marker (Fig. 2M-O). Protein processing in autophagosomes plays a key role in MHC-mediated antigen presentation in DCs, ^63-65^ providing a potential mechanism of enhanced CD4+ T cells induced by N-ETSD in the vaccine construct. Evidence of this T cell interaction with a MoDC infected with N-ETSD translocated to autophagosomes (and, it is assumed, also endosomes and lysosomes) is seen in this phase-contrast microscopy of the N-ETSD and LC3a/b co-labeled cells, which reveals the elongated DC morphology in contrast to the spherical morphology of undifferentiated lymphocytes. Lymphocytes, also distinguished by the absence of infection by hAd5 N-ETSD (lymphocytes lack the hAd5 receptor), were also seen to interact with N-ETSD-expressing MoDCs (Fig. 2U and X).

**Fig. 2.**
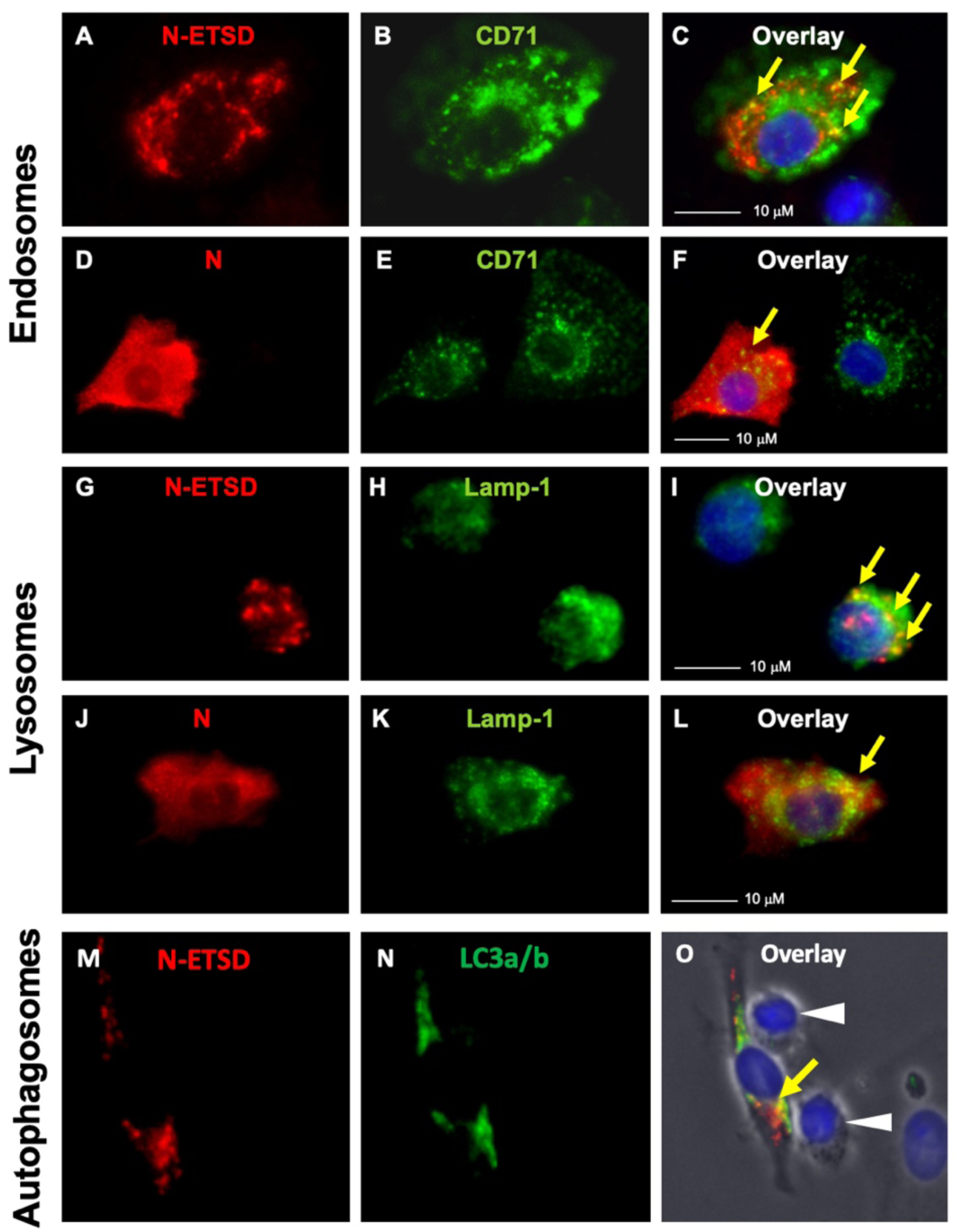
N-ETSD localizes to endosomes, lysosomes, and autophagosomes. MoDCs were infected with Ad5 N-ETSD or N without ETSD and were co-labeled with anti-flag (N, N-ETSD here have a flag tag) and anti-CD71 (endosomal marker), anti-Lamp 1 (lysosomal marker), or anti-LC3a/b antibodies. (A) N-ETSD, (B) CD71, and (C) overlay. (D) N, (E) CD71, and (F) overlay. (G) N-ETSD, (H) Lamp-1, and (I) overlay. (J) N, (K) Lamp-1, and (L) overlay. (M) N-ETSD, (N) LC3a/b, and (O) overlay. N/N-ETSD is red, other markers green, co-localization indicated by yellow arrows, and white arrows indicate lymphocytes.

### Validation of SARS-CoV-2 antibody and cell-mediated immune responses from previously SARS-CoV-2 infected patients and virus-naïve patients for memory T cell recall studies

For the studies described below, plasma samples were collected from four individuals convalescing from SARS-CoV-2 infection as confirmed by antibody assays and patient history as described *Methods*. The presence of anti-Spike IgG, and of neutralizing antibodies by both the cPass ^66^ and live virus assays, were confirmed in all patient samples (Supplementary Fig. S1). Samples were also collected from four virus-naïve individuals and were used as controls.

In additional studies to validate immune responses to SARS-CoV-2 antigens, the binding of previously SARS-CoV-2 infected patient and virus-naïve control individual plasma to human embryonic kidney (HEK) 293T cells transfected with either hAd5 S-Fusion alone or hAd5 S-Fusion + N-ETSD was assessed (Supplementary Fig. S2). This binding reflects the presence of antibodies in plasma that recognize antigens expressed by the hAd5 vectored vaccines. Quantification of histograms showed little or no binding of virus-naïve plasma antibodies to cells expressing either construct, and the highest binding of plasma antibodies from a previously SARS-CoV-2 infected patient to cells expressing the bivalent S-Fusion + N-ETSD construct (Supplementary Fig. S2R). This could be due to either the enhanced cell surface expression of S found in hAd5 S-Fusion + N-ETSD infected HEK 293T cells as compared to hAd5 S-Fusion alone (Supplementary Figs. S3 and S4D) or expression of both S and N antigens.

### N-ETSD optimizes spike antigen expression: binding of plasma antibodies from previously infected SARS-CoV-2 patients is enhanced for hAd5 S-Fusion + N-ETSD infected MoDCs compared to hAd5 S-Fusion or hAd5 S-WT

The studies herein focus on the responses of T cells from previously SARS-CoV-2 infected patients to hAd5 vaccine construct-infected autologous MoDCs. DCs are powerful antigen presenting cells for processing and presenting complex antigens acquired through infection or phagocytosis to elicit a T-cell response. Therefore, in addition to the assessment of binding of patient plasma antibodies to hAd5 vaccine expressing HEK 293T cells, we infected MoDCs from two healthy individuals overnight with hAd5 S-WT, hAd5 S-Fusion, hAd5 S-Fusion + N-ETSD, or hAd5 Null then evaluated gene expression using plasma from a previously SARS-CoV-2 infected patient (Fig. 3A). For both MoDC sources, the highest binding of plasma antibodies from a previously infected patient to the MoDCs was seen after hAd5 S-Fusion + N-ETSD infection (Fig. 3B-D), providing further evidence that antigen expression is optimized in the hAd5 S-Fusion + N-ETSD bivalent vaccine. This finding in a highly relevant *in vitro* system of human MoDCs and plasma is not only an important confirmation of results from testing with HEK 293T cells and commercially-available anti-S RBD antibodies, it represents a potential method to screen plasma for SARS-CoV-2 antigen reactivity.

**Fig. 3.**
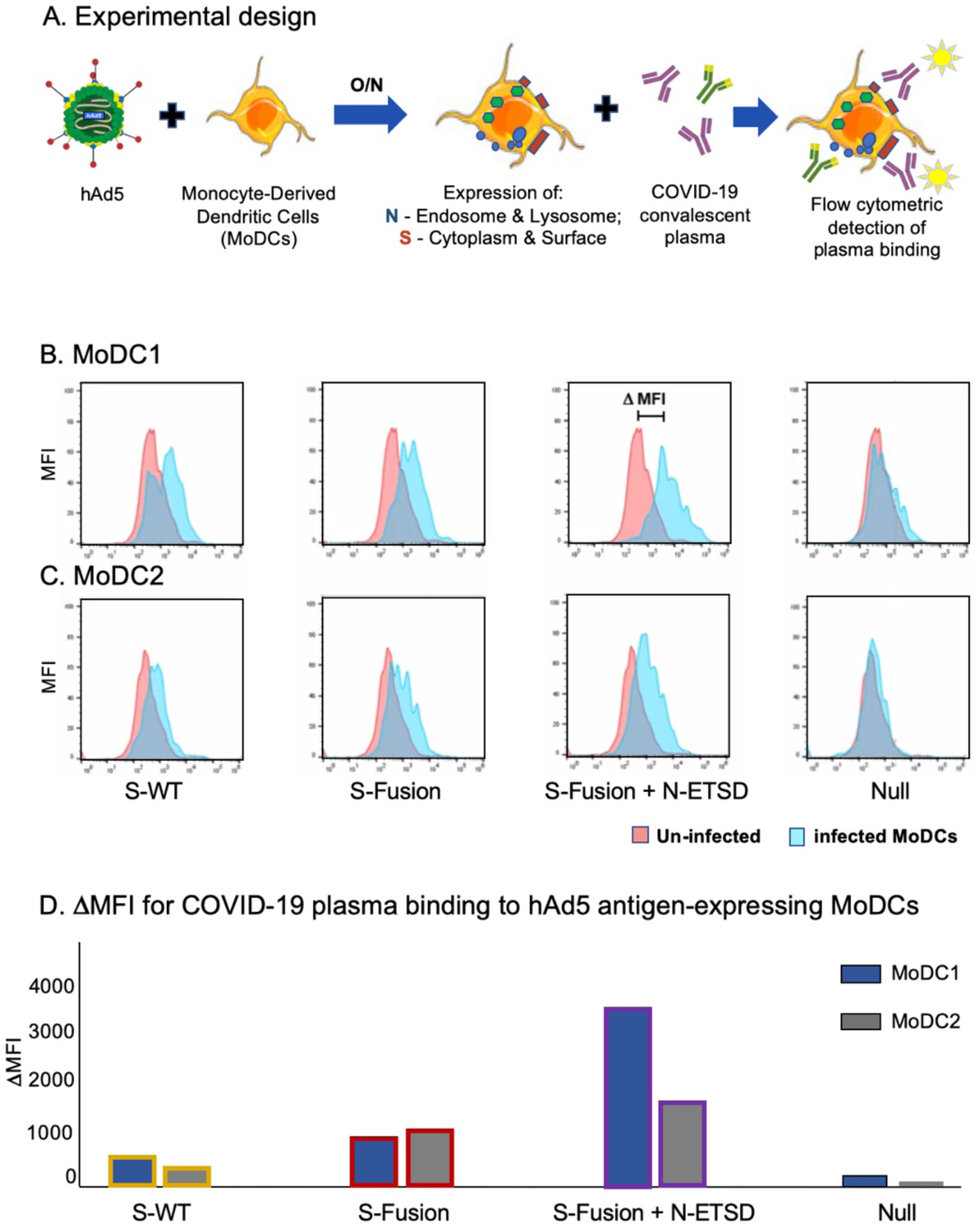
Patient plasma antibodies recognize SARS-CoV-2 antigens expressed by MoDCs after hAd5 S-Fusion + N-ETSD infection. (A) MoDCs from two normal individuals were infected with hAd5 vaccines overnight, then exposed to previously infected patient plasma from a single individual; antibody binding to the DC cell surface was detected by flow cytometry. The flow histograms for hAd5 S-WT, S-Fusion, S-Fusion + N-ETSD, and Null are shown for MoDCs from two sources, (B) MoDC1 and (C) MoDC2. (D) The ΔMFI (difference in MFI for binding to uninfected and infected cells) is graphed for hAd5 S-WT, S-Fusion, S-Fusion + N-ETSD and Null infected MoDCs.

### SARS-CoV-2 peptide pool immune reaction: T cells from previously infected SARS-CoV-2 patients secrete significant levels of interferon-γ (IFN-γ) in response to S1, S2, and N SARS-CoV-2 peptide pools compared to T cells from virus-naïve controls

To demonstrate the reactivity of T cells from four previously infected SARS-CoV-2 patients versus virus-naive T cells from four unexposed individuals, T cells from each group were cultured with autologous MoDCs pulsed with peptide mixes spanning the sequences of N and S proteins. T cells from previously infected SARS-CoV-2 patients but not unexposed subjects secreted IFN-γ in response to SARS-CoV-2 antigens (Fig. 4), validating selective reactivity of T cells from patients previously infected with SARS-CoV-2.

**Fig. 4.**
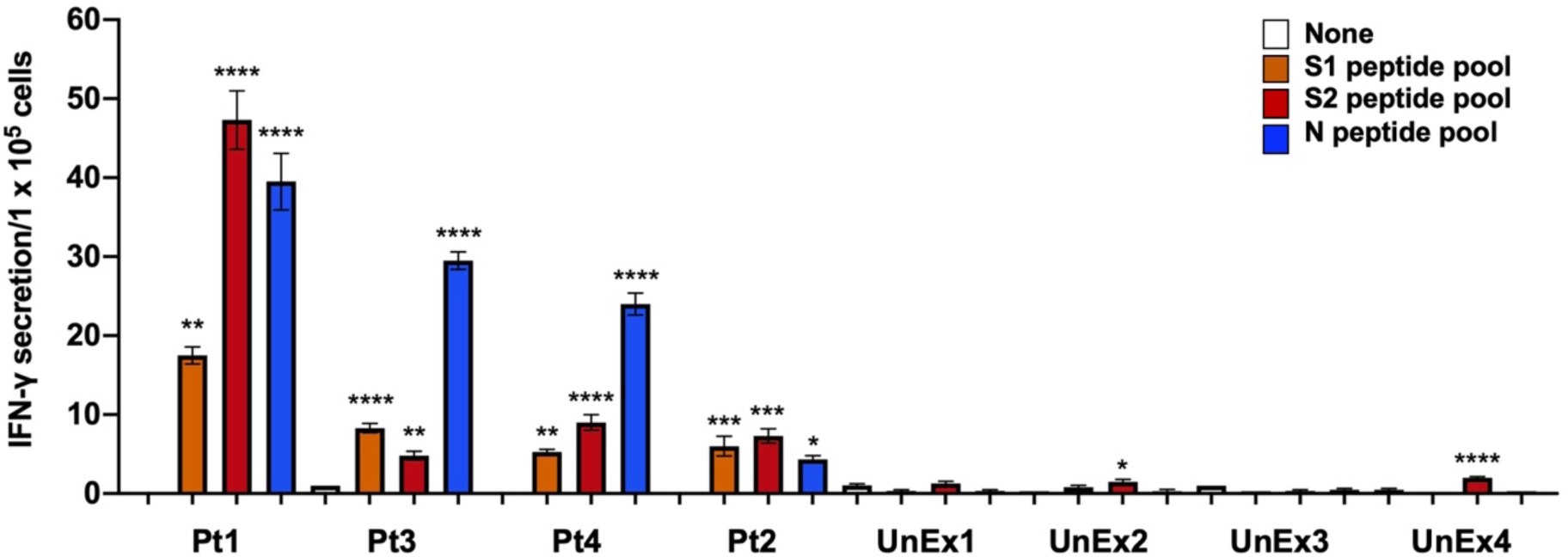
Previously SARS-CoV-2 infected patient and virus-naive T-cell responses to MoDCs pulsed with SARS-CoV-2 peptides. T cells from all four previously SARS-CoV-2 infected patients (Pt) show significant IFN-γ responses to S1, S2, and N peptide pool-pulsed MoDCs as compared to ‘none’. T cells from virus-naïve (unexposed, UnEx) control individuals showed far lower responses. Statistical analysis performed using One-way ANOVA and Tukey’s post-hoc analysis for samples from each patient compared only to ‘none’ where * p<= 0.05, **p<0.01, ***p<0.001, and ****p<0.0001. Data graphed as the mean and SEM; n = 3-4.

### SARS-CoV-2 peptide pool immune reaction: CD4+ T cells from previously infected SARS-CoV-2 patients recognize S and N peptide pool antigens, but CD8+ T cells display greater recognition of N peptide antigens

To confirm S and N antigen recognition by T cells, we assessed IFN-γ responses to SARS-CoV-2 peptide pools by the two major T-cell subpopulations, CD8+ and CD4+ (see selected T-cell phenotypes in Supplementary Fig. S5). CD4+ T cells of the two patient samples tested responded to both the S and N peptide pools, with a higher response to N by Pt3 (Fig. 5B). In contrast, CD8+ T cells from both patients responded to N with high significance, but not to S1 or S2 peptide pools (Fig. 5C and D). These data are consistent with published studies demonstrating T-cell responses against multiple antigens, including S and N in previously infected SARS-CoV-2 patients^37^.

**Fig. 5.**
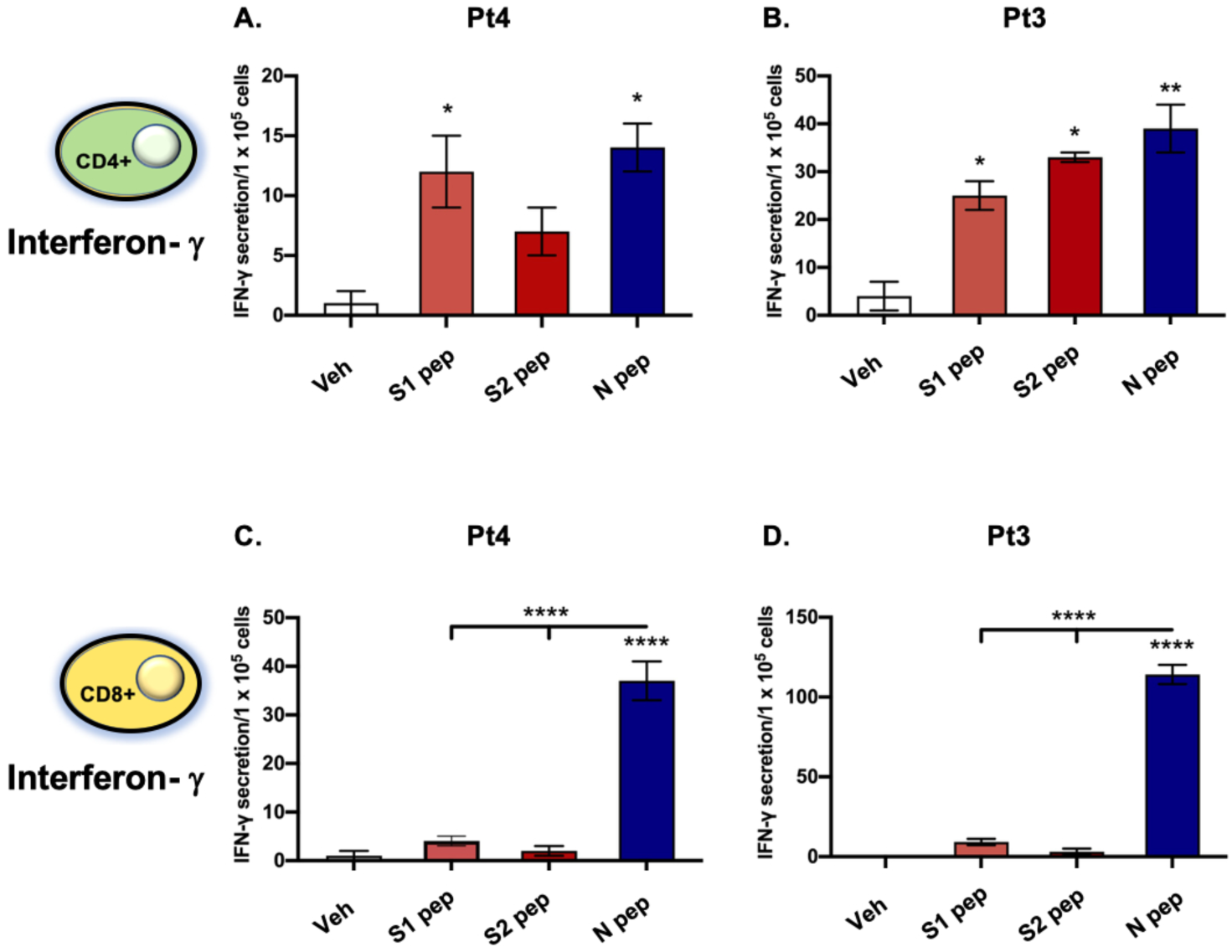
Peptide-pulsed MoDCs from patients previously infected with SARS-CoV-2 stimulate autologous patient T cells to secrete IFN-γ. (A) MoDCs derived from previously infected SARS-CoV-2 patients Pt4 and Pt 3 were pulsed with SARS-CoV-2 peptide mixes (S1, S2 or N) overnight and then incubated with autologous CD4+ (A, B) or CD8+ (C, D) T cells. IFN-γ levels were determined by ELISpot. Statistical analysis performed using One-way ANOVA and Dunnett’s post-hoc multiple comparison analysis to compare each peptide pool to Veh (shown above the bar) or between peptide pools (above line) where *p<0.05, **p<0.01 and ****p<0.00001. Data graphed as mean and SEM; n = 3.

### Autologous MoDCs infected with endo/lysosome-directed nucleocapsid-ETSD elicit higher levels of IFN-γ secretion from CD4+ and CD8+ T cells from previously infected SARS-CoV-2 patients compared to cytoplasmic nucleocapsid protein (hAd5 N)

To evaluate the immune significance of endo/lysosome-localized N-ETSD versus cytoplasmic N, MoDCs were infected with hAd5 constructs (Null, N-ETSD or N) then incubated with autologous CD3+ and CD4+- or CD8+-selected T cells (Fig. 6A). CD3+ T cells from previously infected SARS-CoV-2 patients showed significantly greater IFN-γ secretion in response to N-ETSD than both Null and cytoplasmic N in the two patients where N-ETSD and N were compared (Fig. 6C and D). There were relatively few interleukin-4 (IL-4) secreting CD3+ T cells for all patients (Fig. 6E-G). Both CD4+ and CD8+ selected T-cell populations showed significantly greater IFN-γ responses to N-ETSD than Null (Fig. 6H-M) and in two of three patients, CD4+ and CD8+ T cells showed greater recognition of N-ETSD compared to N. The high IFN-γ and low IL-4 responses indicate a predominant Th1 cytokine response to N/N-ETSD. Cell expression of N-ETSD and N were equivalent in 293T HEK cells (Supplementary Fig. S4G), suggesting the reason for the elevated T-cell response to N-ETSD was likely processing and MHC loading of the epitopes.

**Fig. 6.**
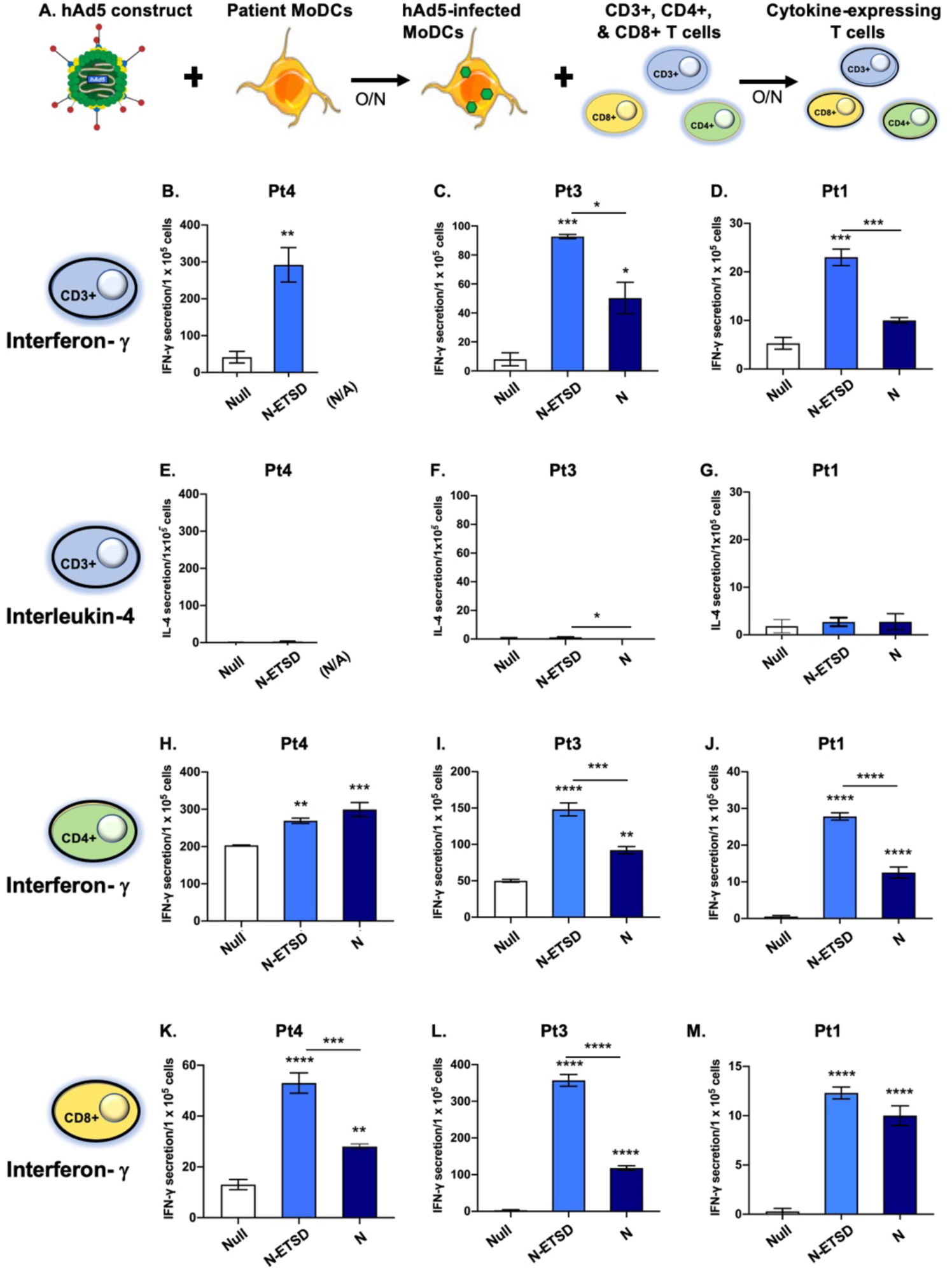
IFN-γ secretion by T cells from previously SARS-CoV-2 infected patients is greater in response to MoDC expression of N-ETSD compared to N. (A) Experimental design. (B-D) Secretion of IFN-γ by autologous CD3+ T cells in response to hAd5-N-ETSD- and hAd5 N-expressing MoDCs is shown. (E-G) Secretion of IL-4 by CD3+ cells in response to infected MoDCs is shown with the same scales as IFN-γ for each. IFN-γ secretion by (H-J) CD4+ and (K-M) CD8+ T cells in response to hAd5-N-ETSD compared to Null are shown. Statistical analysis performed using One-way ANOVA and Tukey’s post-hoc multiple comparison analysis, where *p<=0.05; **p<0.01, ***p<0.001 and ****p<0.0001. Comparison to Null shown above bars, comparison between N-ETSD and N above lines. Data graphed as mean and SEM; n = 3-4.

### Th1 dominant SARS-CoV-2 specific CD4+ and CD8+ memory T-cell recall to nucleocapsid and spike antigens is induced by hAd5 S-Fusion + N-ETSD infection of autologous MoDCs from previously SARS-CoV-2 infected patients

We demonstrated above that N-ETSD is more effective than N in eliciting patient T-cell cytokine responses. We next endeavored to determine whether patients previously infected SARS-CoV-2 generated CD3+, CD4+, CD8+ memory T-cell recall to autologous MoDCs infected with the bivalent vaccine construct hAd5 S-Fusion + N-ETSD. In addition, we compared the individual components of the bivalent vaccine, S-Fusion and N-ETSD, to the bivalent vaccine (S-Fusion + N-ETSD) itself in the same experimental system (Fig. 7A).

**Fig. 7.**
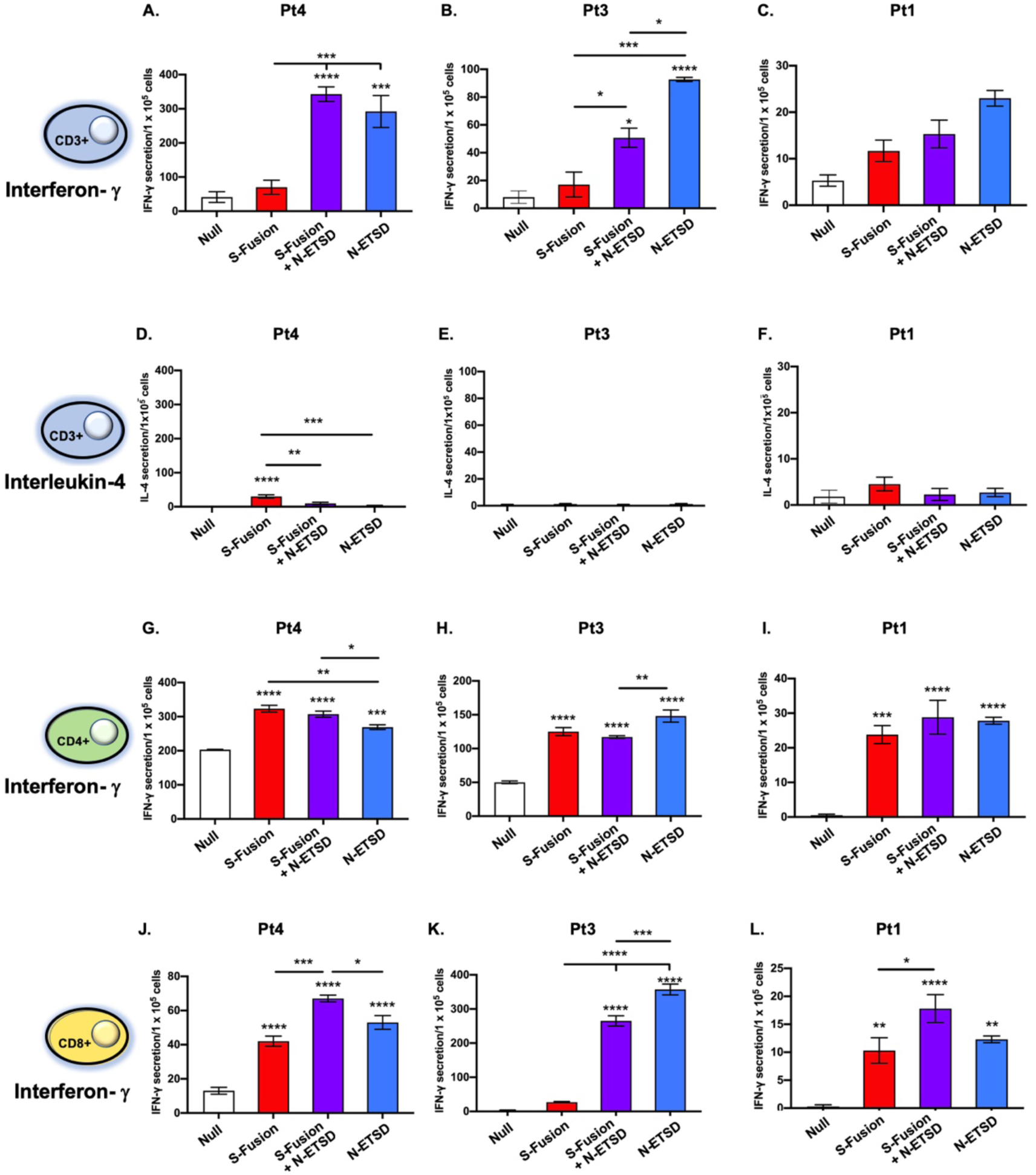
Previously SARS-CoV-2 infected patient T-cell responses to the bivalent vaccine and its individual components reveal distinct antigen specificity of T-cell populations. (A-C) CD3+ T cell IFN-γ responses for three patients. (D-F) CD3+ T cell IL-4 responses. (G-I) CD4+ IFN-γ responses. (J-L) CD8+ IFN-γ responses. Statistical analysis performed using One-way ANOVA and Tukey’s post-hoc multiple comparison analysis to compared each antigen-containing construct to the Null construct, where *p<0.05 and ****p<0.00001. Comparison to Null only above bars; comparison between antigen-expressing vaccines above lines. Data graphed as mean and SEM; n = 3-4.

For total T cells (CD3+), IFN-γ responses were similar for S-Fusion + N-ETSD and N-ETSD with responses to S-Fusion being relatively low (Fig. 7A-C). The number of IL-4 secreting T cells was very low for all (Fig. 7D-F). Based on the increased expression of S in the bivalent vaccine compared to monovalent S-Fusion, the increased T-cell response could be explained by either T cells recognizing increased S or the presence of N. Importantly, these T-cell responses were characterized by a predominance of IFN-γ (Th1) relative to IL-4 (Th2).

CD4+ T cells from all three patients showed significantly greater recognition of all three constructs compared to Null (Fig. 7G-I). While there were greater responses to specific constructs in some individuals, overall the responses to S-Fusion, S-Fusion + N-ETSD and N-ETSD were similar. CD8+ T cells from all three patients recognized the bivalent and N-ETSD vaccines at a significantly higher level than Null; in only two of three patients did CD8+T cells recognize S-Fusion to a significant degree above Null (Fig. 7J-L). These data indicate that T cells from previously infected SARS-CoV-2 patients have reactivity and immune memory recall to both of the vaccine antigens (S and N) in our vaccine vector.

## DISCUSSION

We previously showed that our hAd5 S-Fusion + N-ETSD vaccine elicits both T-cell immunity and neutralizing antibodies in a murine pre-clinical model. ^12^ Here we demonstrate the CD4+ and CD8+ memory T cells of previously infected SARS-CoV-2 patients, but not unexposed individuals, recall SARS-CoV-2 antigens expressed by hAd5 S-Fusion - N-ETSD infected dendritic cells. These data support the hypothesis that SARS-CoV-2 antigens delivered to cells by the next generation hAd5 platform are expressed in human cells in a conformationally relevant manner and available to elicit an adaptive immune response, critical for vaccine efficacy.

One feature of S-Fusion is the higher expression of S RBD compared to S-WT, confirmed here ^12^ by infection of human cells. This was a goal of our vaccine design based on findings from earlier studies of S cryo-electron micrograph structures that suggested RBD epitopes would be largely unavailable for immune detection ^24,25^ We gain a further advantage by combining S with N in the bivalent vaccine, through the ability of N to enhance immune detection of S, a phenomenon that has been observed by others for gene expression in general. ^67^

We designed the vaccine-expressed N protein to traffic to the endosomal/lysosomal subcellular compartments, a key antigen presenting pathway to stimulate CD4+ T cells so that they can license dendritic cells to activate naïve CD8+ CTL. ^68,69^ Here, we confirmed N-ETSD does localize to endosomes and lysosomes, as well as to autophagosomes, in MoDCs. Both endosomal and lysosomal targeting are desirable for enhanced antigen presentation and CD4+ T-cell activation. ^70,71^ Lysosomes can fuse acidic autophagosomes,^72^ facilitating protein processing; this has important implications for effective immune stimulation by modulation of MHC class II presentation. ^62^ We confirmed this desired effect by demonstrating that the T cells of previously infected SARS-CoV-2 patients more readily recognized N-ETSD than N. The findings of Gupta *et al*. ^71,73^ and the data presented here strongly support the potential of enhanced efficacy of a vaccine construct specifically expressing the modified N-ETSD.

T cells are critical for elimination of SARS-CoV infection. ^36,74-78^ Here, hAd5 expressed S and N elicited strong antigen-specific IFN-γ, but virtually no IL-4 secretion from T cells of previously infected SARS-CoV-2 patients, pointing to Th1 dominance. Antiviral Th1 cytokine responses eliminate a variety of viruses from infected hosts ^79^ including the virus closely related to SARS-CoV-2, SARS-CoV. ^80^ These data are also consistent with our studies in preclinical models. ^12^ Importantly, our data suggests that both S and N are targets of CD4+ T cells that help both antibody production from B cells ^81-83^ and CD8+ T cell memory, which together function to kill virus-infected targets. ^84^ The recognition of these vaccine antigens by the T-cell subsets are consistent with immune control of the pathogen. Intriguingly, our hAd5 S-Fusion + N-ETSD T-cell biased vaccine has the potential to not only provide protection for uninfected patients, but also to be utilized as a therapeutic for already infected patients to induce rapid clearance of the virus by activating T cells to kill the virus-infected cells, thereby reducing viral replication and lateral transmission.

Importantly, the T cell recall of N-ETSD was shown to be Th1 dominant as shown by the vigorous interferon-γ response and the low IL-4 response.

We believe the results of our studies, particularly in light of reports of N-driven T-cell responses in asymptomatic and mild COVID patients, ^2,37^ suggest that N is an important contributor to immunity in humans and its presence in a vaccine increases the opportunity for effective protection against COVID-19. The critical role of the T-cell response - specifically a CD8+ T-cell response to N - was brought to the forefront by Peng *et al*. ^85^ who compared these responses for mild or severe COVID-19 cases (with unexposed controls). In mild cases, N- and (M-) specific CD8+ cell responses predominated, suggesting T-cell responses to these antigens might be vital to prevent progression of disease and concluded non-spike antigens be part of a vaccine for COVID-19. The work presented here and earlier ^12^ support this hypothesis.

The display of immune recall reported here, together with our other findings, strongly supports the ongoing testing of the bivalent S-Fusion + N-ETSD in human clinical trials to study the efficacy and safety of the vaccine and to assess its potential to provide robust, durable immune protection against SARS-CoV-2 infection and COVID-19.

## METHODS

### The hAd5 [E1-, E2b-, E3-] platform and constructs

For studies herein, the 2^nd^ generation hAd5 [E1-, E2b-, E3-] vector was used (Fig. 1A) to create viral vaccine candidate constructs. hAd5 [E1-, E2b-, E3-] backbones containing SARS-CoV-2 antigen expressing inserts and virus particles were produced as previously described.^17^ In brief, high titer adenoviral stocks were generated by serial propagation in the E1- and E2b-expressing E.C7 packaging cell line, followed by CsCl_2_ purification, and dialysis into storage buffer (2.5% glycerol, 20 mM Tris pH 8, 25 mM NaCl) by ViraQuest Inc. (North Liberty, IA). Viral particle counts were determined by sodium dodecyl sulfate disruption and spectrophotometry at 260 and 280 nm. Viral titers were determined using the Adeno-X™ Rapid Titer Kit (Takara Bio). The constructs created included:

i. S-WT: S protein comprising 1273 amino acids and all S domains: extracellular (1-1213), transmembrane (1214-1234), and cytoplasmic (1235-1273) (Unitprot P0DTC2);
ii. S Fusion: S optimized to enhance surface expression and display of RBD;
iii. N: Nucleocapsid (N) wild type sequence protein containing tags for immune detection;
iv. N-ETSD: N with an Enhanced T-cell Stimulation Domain (ETSD) together with tags for immune detection; and
v. Bivalent S-Fusion + N-ETSD;

### Collection of plasma and peripheral blood mononuclear cells from patients with confirmed previous SARS-CoV-2 infection and from virus-naïve volunteers

Blood was collected with informed consent via venipuncture from volunteers who had either not been exposed (UNEX) to SARS-CoV-2 as confirmed by ELISA and multiple negative SARS-CoV-2 tests or who had recovered from COVID-19 as indicated by recent medical history and a positive SARS-CoV-2 antibody test (Patients, Pt). A third source of whole blood was apheresis of healthy subjects from a commercial source (HemaCare). Peripheral blood mononuclear cells (PBMCs) were isolated from whole blood by density gradient centrifugation and plasma was collected after density gradient centrifugation.

Monocyte-derived dendritic cells (MoDC) were differentiated from PBMC using GM-CSF (200U/ml) and IL-4 (100U/ml) as previously described^86^. Briefly, monocytes were enriched by adherence on plastic, while the non-adherent cells were saved and frozen as a source of lymphocytes, specifically T cells. Adherent cells were differentiated into dendritic cells (3-5 d in RPMI containing 10% FBS), then frozen in liquid nitrogen for later use. T cells were enriched from the non-adherent fraction of PBMC using MojoSort (BioLegend CD3 enrichment). CD4+ and CD8+ T cells were enriched using analogous kits from the same manufacturer. Efficiency of the cell separations was evaluated by flow cytometry (Supplementary Fig. S5).

### Infection of MoDCs with hAd5 N-WT or N-ETSD and labeling with anti-N, anti-CD71, anti-LAMP-1, and Anti-LC3a/b antibodies

Freshly thawed MoDCs were plated on 4-well Lab-Tek II CC2 Chamber Slides, using 3 ×10^4^ cells per well and transduction performed at MOI 5000 one hour after plating using hAd5 N-ETSD or hAd5 N. Slides were incubated o/n at 37°C, fixed in 4% paraformaldehyde for 15 minutes, then permeabilized with 1% Triton X100, in PBS) for 15 min. at room temperature. To label N, cells were then incubated with an anti-flag monoclonal (Anti-Flag M2 produced in mouse) antibody at 1:1000 in phosphate buffered saline (PBS) with 3% BSA, 0.5% Triton X100 and 0.01% saponin overnight at 4°C, followed by three washes in PBS and a 1 hour incubation with a goat anti-Mouse IgG (H+L) Highly Cross-Adsorbed Secondary Antibody, Alexa Fluor Plus 555 (Life Technologies) at 1:500. For co-localization studies, cells were also incubated overnight at 4°C with a rabbit anti-CD71 (transferrin receptor, recycling/sorting endosomal marker) antibody (ThermoFisher) at 1:200; sheep anti-Lamp1 Alexa Fluor 488-conjugated (lysosomal marker) antibody (R&D systems) at 1:10; or a rabbit monoclonal anti human LC3a/b (Light Chain 3, autophagy marker) antibody (Cell Signaling Tech #12741S) used at 1:100. After removal of the primary antibody, two washes in PBS and three washes in PBS with 3% BSA, cells were incubated with fluor-conjugated secondary antibodies when applicable at 1:500 (Goat anti-Rabbit IgG (H+L) secondary antibody, Alexa Fluor 488; 1:500 dilution) for 1 hour at room temperature. After brief washing, cells were mounted with Vectashield Antifade mounting medium with DAPI (Fisher Scientific) and immediately imaged using a Keyence all-in-one Fluorescence microscope camera and Keyence software.

### Binding of plasma antibodies from previously SARS-CoV-2 infected patients to antigens expressed by vaccine-infected MoDCs

Binding of plasma antibodies from previously infected subjects to antigens expressed on the surface of MoDCs was determined by differentiation of MoDCs from peripheral blood mononuclear cells (PBMC) to DC, infection of the MoDCs, incubation with previously infected patient plasma, and detection of binding to infected and uninfected MoDCs by flow cytometry.

MoDCs were infected (0.5 × 10^6^/well in 12 well plates) at MOI 5000 using hAd5 S-WT, S-Fusion, S-Fusion + N-ETSD or a ‘Null’ construct that expresses green fluorescent protein (GFP). One day after infection, the MoDCs were detached using EDTA (0.5 mM), gently pipetted and transferred for incubation with previously infected patient plasma at 1:100 dilution from a single patient (Pt4) at (4°C) for 30 minutes, and plasma antibodies detected on the MoDC surface by goat anti-human IgG (phycoerythrin conjugated). Cells were acquired as described above for flow cytometric analyses. Data were graphed as the ΔMFI, that is, the difference in binding between infected and uninfected MoDCs.

### Previously SARS-CoV-2 infected patient and virus-naïve individual T cell and selected CD4+ and CD8+ T cell secretion of IFN-γ in response to MoDCs pulsed with SARS-CoV-2 peptide antigen pools

The ability of T cells from previously infected patients used in these studies to recognize SARS-CoV-2 antigens *in vitro* was validated and then similar analyses were performed for selected CD4+ and CD8+ T cells. Briefly, MoDC (2 × 10^4^) were pulsed with SARS-CoV-2 peptide antigens (1μg/ml, PepMix S comprising the S1 and S2 pools PM-WCPV-S-1; and N PM-WCPV-NCAP-1, both JPT Peptide Technologies) then autologous T cells (1 × 10^5^), enriched from the non-adherent fraction of PBMC using MojoSort (BioLegend CD3 enrichment) were added in enriched RPMI (10% human AB serum). Cells were cultured in a microtiter plate (Millipore) containing an immobilized primary antibody to target IFN-γ, overnight (37°C), then IFN-γ spot forming cells enumerated by ELISpot. For ELISpot detection, after aspiration and washing to remove cells and media, IFN-γ was detected by a secondary antibody to cytokine conjugated to biotin. A streptavidin/horseradish peroxidase conjugate was used detect the biotin-conjugated secondary antibody. The number of spots per well (1 × 10^5^ cells), was counted using an ELISpot plate reader. IL-4 was measured by ELISpot using a kit (MabTech) with wells precoated with anti-IL-4 antibody and following the manufacturer’s instructions. Remaining steps for IL-4 detection were identical to those for IFN-γ, but with alkaline phosphatase detection rather than peroxidase.

### Determination of previously SARS-CoV-2 infected patient-derived T-cell reactivity in response to autologous hAd5 vaccine-infected MoDCs

MoDCs were infected with hAd5 S-Fusion, S-Fusion + N-ETSD, N-ETSD, N or GFP/Null constructs and incubated overnight at 37°C. The infected MoDCs were cultured with CD3+, CD4+, or CD8+ T-cells from the same individuals overnight. Antigen specific T-cell responses were enumerated using ELISpot as described above.

## Data Availability

All relevant data are presented in the manuscript

## SUPPLEMENTARY MATERIALS

### Supplementary Methods

#### Quantification of anti-SARS-CoV-2 spike antibodies in plasma from patients previously infected with SARS-CoV-2

The presence of anti-SARS-CoV-2 spike antigen antibodies in previously infected patient plasma used in studies here was validated. IgG against SARS-CoV-2 spike was detected in plasma using ELISA. Briefly, EIA/RIA plates were coated with a solution of purified recombinant SARS-CoV-2-derived Spike protein (1μg/ml, S-Fusion. ImmunityBio, Inc.) suspended in coating buffer (0.05 M carbonate-bicarbonate, pH 9.6) and incubated overnight at 4°C. Plates were washed three times with TPBS solution (PBS + 0.05% Tween 20). Blocking solution (2% non-fat milk in TPBS) was added and incubated at room temperature (RT, 1h). Serial dilutions of plasma were prepared in 1% non-fat milk in TPBS. Plates were washed as described above and serial dilutions of plasma were added to the plate and incubated at RT (1h). Plates were washed three times. Goat anti-Human IgG (H+L) HRP-conjugated secondary antibody (1:6000 dilution) prepared in 1% non-fat milk/TPBS was added and incubated at RT (1h). Plates were washed three times. Substrate (3,3’,5,5’-tetramethylbenzidine (TMB)) was added to each well and incubated at RT (10min). The reaction was stopped by addition of sulfuric acid (1N H_2_SO_4_). The optical density (450 nm) was measured by a Synergy 2 (BioTek Instruments, Inc.) plate reader. Data were analyzed using Prism 8 (GraphPad Software, LLC).

#### cPass™ Neutralizing Antibody Detection

The GenScript cPass™ (https://www.genscript.com/cpass-sars-cov-2-neutralization-antibody-detection-Kit.html) kit for detection of neutralizing antibodies was used according to the manufacturer’s instructions.^66^ The kit detects circulating neutralizing antibodies against SARS-CoV-2 that block the interaction between the S RBD with the ACE2 cell surface receptor. It is suitable for all antibody isotypes.

To evaluate the levels of anti-SARS-CoV-2 neutralizing antibodies in plasma, dilutions of plasma were incubated with horseradish peroxidase-conjugated spike RBD (37°C, 30min). The RBD-plasma mixture was added to a microtiter plate coated with ACE2 and incubated at 37°C (15 min). Plates were washed and substrate (TMB) added at room temperature (15 min). The reaction was stopped and plates read on a plate reader at 450 nm.

#### Live virus assay of plasma neutralization of infection

The ability of previously SARS-CoV-2 infected patient plasma used in studies here to neutralize SARS-CoV-2 infection *in vitro* was also validated. All aspects of the assay utilizing virus were performed in a BSL3 containment facility according to the ISMMS Conventional Biocontainment Facility SOPs for SARS-CoV-2 cell culture studies. Vero E6 kidney epithelial cells from *Cercopithecus aethiops* (ATCC CRL-1586) were plated (20,000 cells/well) in a 96-well format and 24 hours later, cells were incubated with antibodies or heat inactivated plasma previously serially diluted in 3-fold steps in DMEM containing 2% FBS, 1% NEAAs, and 1% Pen-Strep; the diluted samples were mixed 1:1 with SARS-CoV-2 in DMEM containing 2% FBS, 1% NEAAs, and 1% Pen-Strep at 10,000 TCID 50/mL for 1 hr at 37°C, 5% CO_2_. This incubation did not include cells to allow for neutralizing activity to occur prior to infection. For detection of neutralization, the virus/sample mixture (120 μL) was transferred to the Vero E6 cells and incubated (48 hours) before fixation with 4% PFA. Each well received virus (60 μl) or an infectious dose of 600 TCID50. Control wells, including six wells on each plate for no virus and virus-only controls, were used. The percent neutralization was calculated as 100-((sample of interest-[average of “no virus”])/[average of “virus only”])*100) with a stain for CoV-2 Np imaged on a Celigo Imaging Cytometer (Nexcelom Bioscience).

#### Transfection of HEK 293T cells for the study of previously SARS-CoV-2 infected patient plasma antibody binding

To assess binding of plasma from a previously infected patient to antigens expressed by hAd5 S-Fusion and hAd5 S-Fusion + N-ETSD vaccines, plasma was incubated with construct-transfected HEK 293T cells and binding determined by flow cytometry. HEK 293T cells (2.5 × 10^5^ cells/well in 24 well plates) were grown in DMEM (Gibco) with 10% FBS and 1X PSA (100 units/mL penicillin, 100 μg/mL streptomycin, 0.25 μg/mL Amphotericin B) at 37°C. Cells were either left untransfected or were transfected with 0.5 µg of S-Fusion or S-Fusion + N-ETSD hAd5 plasmid DNA using a JetPrime transfection reagent (Polyplus) according to the manufacturer’s instructions. Twenty-four hours later, cells were incubated for 30 min. with previously infected patient or healthy (unexposed, UnEx) plasma that had been serially diluted 10-fold for 30 min. Plasma IgG was labeled using a goat anti-human IgG-phycoerythrin conjugated and labeled cells were acquired using the Thermo-Fisher Attune NxT flow cytometer and analyzed using FlowJo Software to determine Mean Fluorescence Intensity (MFI) values of both the untransfected and transfected cells. Results were graphed as the difference in MFI between untransfected and transfected cells for the S-Fusion and S-Fusion + N-ETSD constructs over the plasma dilutions.

#### Spike (S) receptor binding domain (RBD) expression in hAd5 S-WT, S-Fusion and S-Fusion + N-ETSD infected HEK 293T cells

Relative levels of cell-surface expression of S RBD after infection of human embryonic kidney (HEK) 293T cells with various S-expressing hAd5 vaccines was determined by flow cytometry. HEK 293T cells (2.5 × 10^5^ cells/well in 24 well plates) were grown in DMEM containing 10% FBS and PSA (100 units/mL penicillin, 100 μg/mL streptomycin, 0.25 μg/mL Amphotericin B) at 37°C. Cells were either uninfected or infected with hAd5 S-WT, S-Fusion, or S-Fusion + N-ETSD viral particles at a multiplicity of infection (MOI) of 10. For detection of spike on Days 1, 2, 3 and 7 after infection, cells were transferred by gently pipetting into medium and labeled with an anti-RBD monoclonal antibody (clone D003 Sino Biological) and F(ab’)2-Goat anti-Human IgG-Fc secondary antibody conjugated with R-phycoerythrin (Thermofisher). Labeled cells were acquired using a Thermo-Fisher Attune NxT flow cytometer and analyzed using Flowjo Software.

#### Immunoblots for full-length spike and nucleocapsid

HEK 293T cells were cultured, transfected and underwent flow cytometric analysis as described in *Methods* in the main text, but were probed with an Anti-Spike S2 (Sino Biological) antibody. Cells similarly transfected were harvested three days after transfection in RIPA lysis buffer with 1X final Protease Inhibitor cocktail (150 μl). After protein assay, equivalent amounts of protein were run on a 4 to 12% gradient polyacrylamide gel (BioRad) and transferred to nitrocellulose membranes using semi-dry transfer apparatus. Anti-Spike S2 (Sino Biological) and anti-FLAG (for flag-tagged N) (Sigma-Aldrich) antibodies were used as primary antibodies followed by labeling with the appropriate secondary antibody using the Ibind Flex platform. Antibody-specific signals were detected with an infrared Licor Odyssey instrument.

#### Co-transfection of GFP and hAd5 plasmids

HEK 293T cells were transfected using JetPrime reagent as described in main text *Methods* with GFP-expressing plasmids in combination with a null plasmid, N, or N-ETSD; all at 250 ng for each (total of 500 ng). After 24 hours, GFP expression was evaluated by flow cytometry.

### Supplementary Results

#### Confirmation of anti-spike IgG and neutralizing antibodies by cPass and live virus assays of plasma from four patients previously infected with SARS-CoV-2

In experiments described below, plasma from patients (Pt) with confirmed previous SARS-CoV-2 infection was used to support our hypothesis that hAd5 vaccine antigens are similar to antigens presented during natural infection and that antibodies raised in response to exposure of the immune system to such antigens will be neutralizing. It was therefore important for us to confirm the Pt plasma samples that were to be used for binding studies contain sufficient levels of anti-spike antibodies and that these antibodies have the ability to neutralize infection. The anti-spike IgG levels in plasma are shown in Fig. S1A, distinguishing the plasma of previously infected SARS-CoV-2 patients from the unexposed subjects. In two independent neutralization assays, plasma from four previously infected SARS-CoV-2 patients neutralized S-RBD binding to ACE-2 in the cPass assay (Fig. S1B) and live virus infection (Fig. S1C). All four plasma samples used in subsequent studies neutralized viral infection, whereas plasma from individuals unexposed (UnEx) to the virus to be used as controls in the studies below did not.

**Fig S1.**
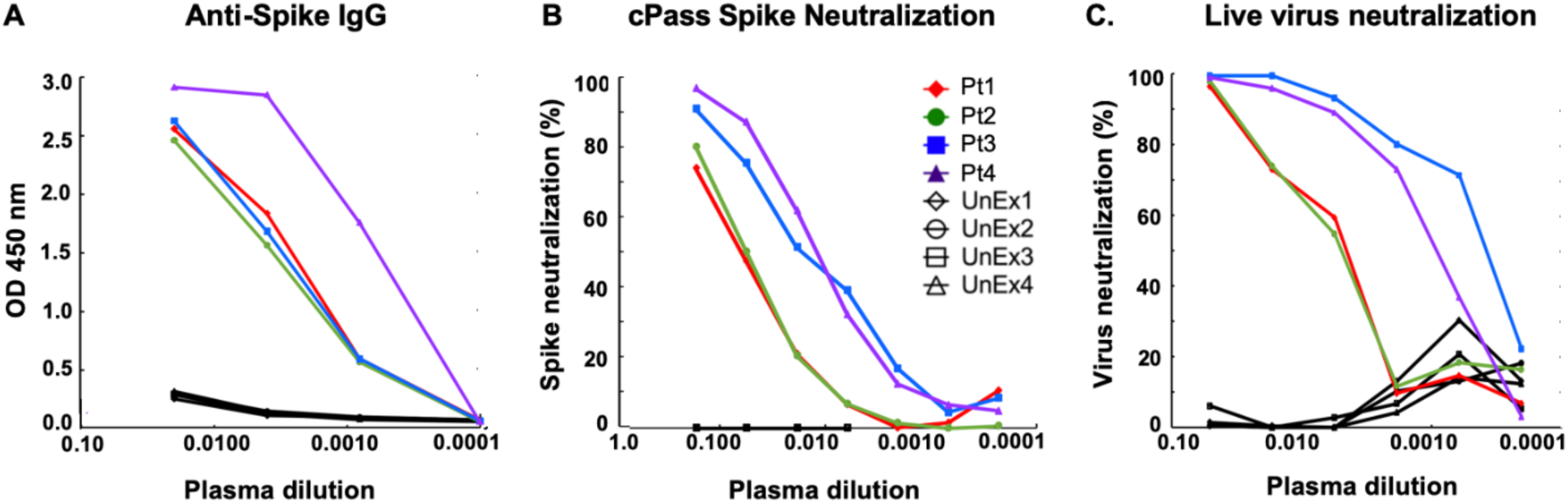
Plasma samples from patients (Pt) previously infected with SARS-CoV-2 contain neutralizing antibodies. (A) The levels of anti-spike IgG as determined by OD at 450 nm are shown. (B) All four Pt plasma samples contained antibodies that blocked RBD-ACE2 binding and contained SARS-CoV-2 neutralizing antibodies; Pt3 and Pt4 were particularly potent.

#### Binding of plasma from patients previously infected with SARS-CoV-2 to hAd5 S-Fusion + N-ETSD transfected HEK 293T cells is enhanced as compared to hAd5 S-Fusion transfected cells

We hypothesized that antibodies from the plasma of previously infected SARS-CoV-2 patients, but not unexposed individuals, would detect expression of hAd5 infected human cells. Plasma from four previously infected SARS-CoV-2 patients and from four healthy, UnEx controls were incubated with HEK 293T cells infected with either hAd5 S-Fusion or hAd5 S-Fusion + N-ETSD and binding detected by flow cytometry. For both hAd5 S-Fusion (Fig. S2A-D) and hAd5 S-Fusion + N-ETSD (Fig. S2E-F), binding by UnEx plasma was almost undetectable or low, whereas binding by plasma from previously infected patients was detectable for both hAd5 S-Fusion (Fig. S2I-L) and hAd5 S-Fusion + N-ETSD (Fig. S2M-P). When binding is represented by the difference in mean fluorescence intensity (ΔMFI) and graphed by increasing fold dilution, it reveals that plasma from previously infected patients shows much higher binding to hAd5 S-Fusion + N-ETSD infected cells, particularly at the 1:10 and 1:100 dilutions (Fig. S2R). While the target of the antibodies in plasma from previously infected patients in this experimental system could be S or N, the greatest likelihood is S, because S antigens are displayed on the cell surface whereas N protein are directed to an endosomal/lysosomal (and as shown in the main text, autophagosomes) compartments. However, some N cell-surface display cannot be excluded. Interestingly, Pt3 and Pt4, shown to have potent neutralizing capability (Fig. S1C), show the highest binding to S-Fusion + N-ETSD transfected cells here. The study is an important confirmation that expression of S is enhanced when in the context of the bivalent vaccine and that it is in conformationally relevant form that can be recognized by antibodies from SARS-CoV-2 infected persons in response to natural infection.

**Fig S2.**
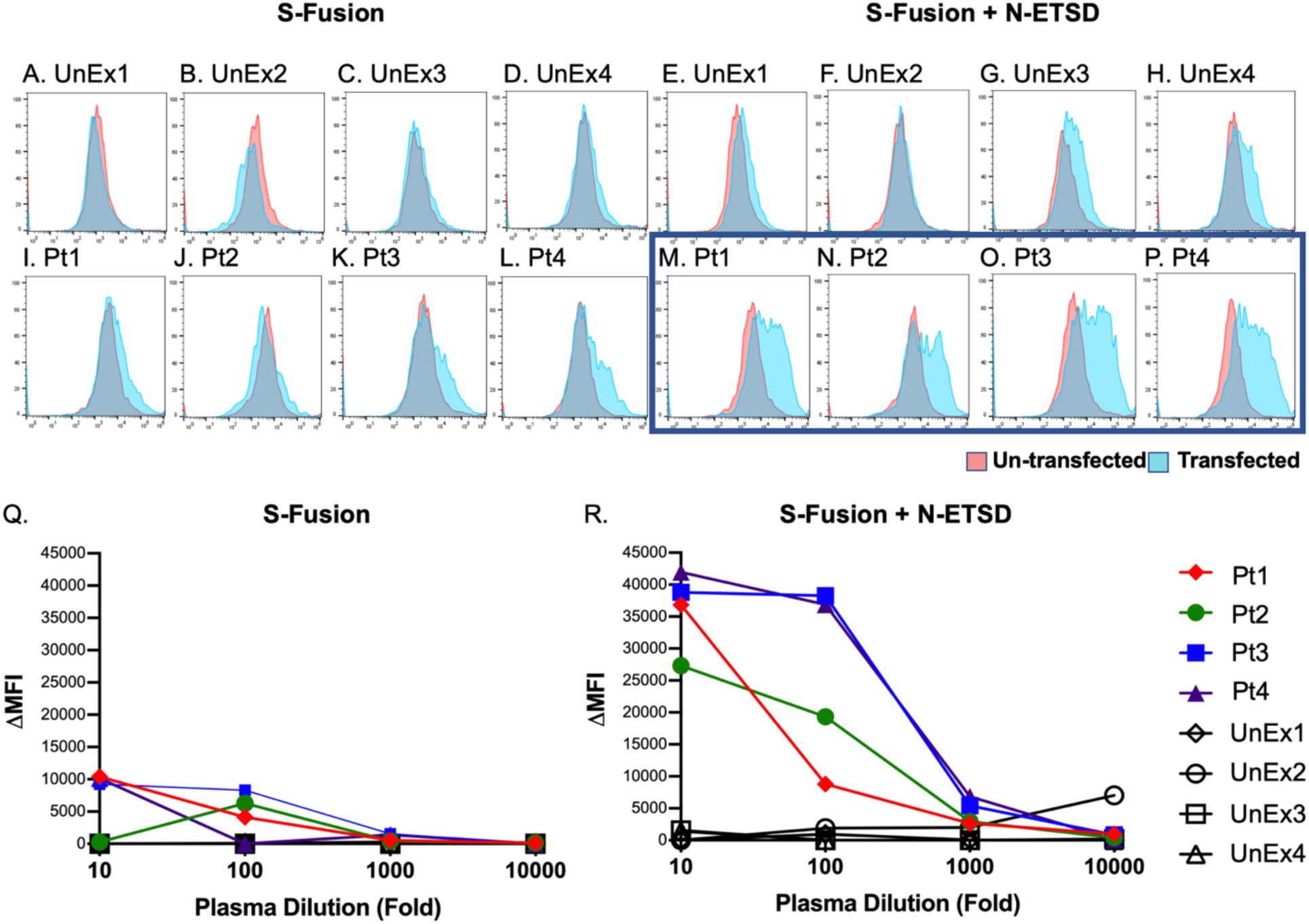
Plasma from previously SARS-CoV-2 infected patients shows greater binding to S-Fusion + N-ETSD surface antigens compared to S-Fusion alone after transfection of HEK 293T cells. HEK 293T cells that were either uninfected (pink) or infected (blue) with S-Fusion alone or bivalent S-Fusion + N-ETSD were exposed to plasma from either unexposed (UnEx) controls or previously infected SARS-CoV-2 patients (Pt). Flow histograms for (A-D) UnEx with S-Fusion; (E-H) UnEx with S-Fusion + N-ETSD; (I-L) Pt with S-Fusion; and (M-P) Pt with S-Fusion + N-ETSD, all n = 4, are shown. When the ΔMFI (difference in binding to transfected versus untransfected cells) is graphed, it is apparent that compared to (Q) S-Fusion, (R) binding of Pt plasma to S-Fusion + N-ETSD transfected cells is much higher and that plasma from UnEx individuals shows little binding.

#### hAd5 S-Fusion + N-ETSD infection of HEK 293T cells leads to enhanced cell-surface S RBD expression compared to hAd5 S-WT or hAd5 S-Fusion alone

In our initial report on the hAd5 S-Fusion + N-ETSD vaccine, ^12^ we showed that after *transfection* of human embryonic kidney (HEK) 293T cells, higher cell surface expression of S RBD is seen with S-Fusion + N-ETSD compared to S-WT or S-Fusion alone. When the vaccine is delivered *in vivo*, however, genes encoding the antigens will be expressed after *infection* by the hAd5 [E1-, E2b-, E3-] vector. Here we show RBD cell surface enhancement of expression with S-Fusion + N-ETSD after infection (Fig. S3C). S RBD is highest one day after infection of HEK 293T cells with hAd5 S-Fusion + N-ETSD. This cell-surface expression is higher than with hAd5 S-Fusion alone (Fig. S3B) and expression with hAd5 S-WT is nearly undetectable (Fig. S3A). Optimizing cell-surface expression results in increased immune cell responses to vaccine antigens, a primary goal of our vaccine design efforts.

**Fig S3.**
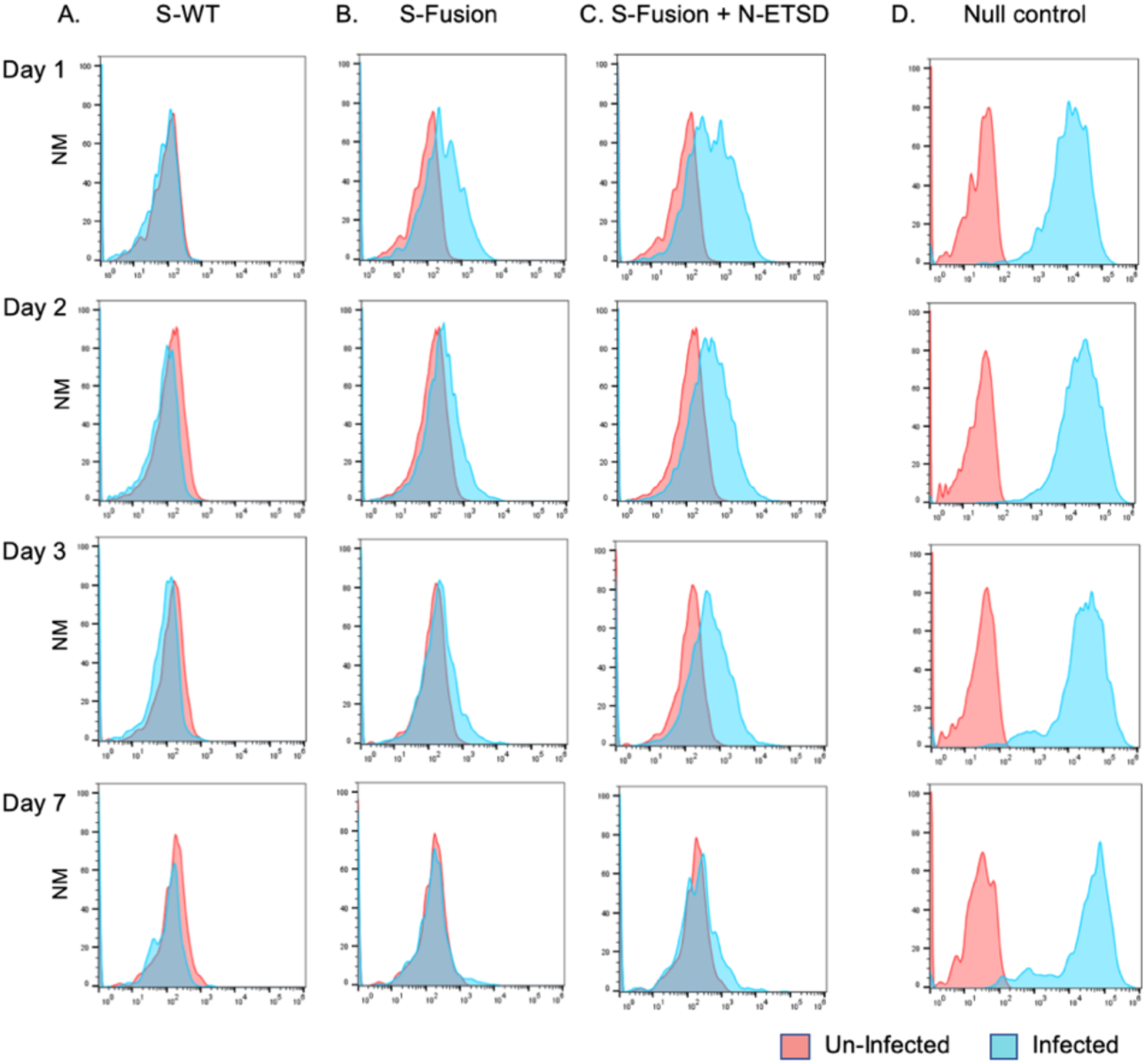
Cell-surface expression of spike (S) RBD is increased after infection of HEK 293T cells with hAd5 S-Fusion and hAd5 S-Fusion + N-ETSD. Surface expression of S RBD as detected by an S RBD-specific antibody (Sino Biological clone D003) is very low after infection of HEK cells with (A) hAd5 S-WT; surface expression is increased with (B) hAd5 S-Fusion, but is the highest for (C) hAd5 S-Fusion + N-ETSD. The increase was greatest on Day 1 of the time course on which flow cytometric analysis was performed. The difference in antibody binding is represented by the increased area of signal from infected cells (blue) compared to uninfected cells (pink), normalized to mode (NM). The null control is hAd5 GFP.

#### S expression enhanced when co-expressed with N-ETSD compared to S-Fusion alone and GFP expression is also enhanced when co-expressed with N-ETSD

To determine whether addition of N to the vaccine construct improved expression of S epitopes in addition to the RBD, we probed hAd5 S-Fusion + N-ETSD transfected HEK 293T cells with an anti-S2 antibody (Fig. S4B) and then performed an immunoblot using the anti-S2 antibody. We observed increased S after transfection with S-Fusion + N-ETSD as compared to S-WT or S-Fusion (Fig. S4D). Expression of N and N-ETSD are similar (Fig. S4G) and expression of N-ETSD is not increased by co-expression with S-Fusion as compared to S-WT (Fig. S4H). This enhancement of protein expression by N-ETSD is not limited to S; GFP expression is also increased when it is co-expressed with N-ETSD as compared to Null or N (Fig. S4I-K).

**Fig S4.**
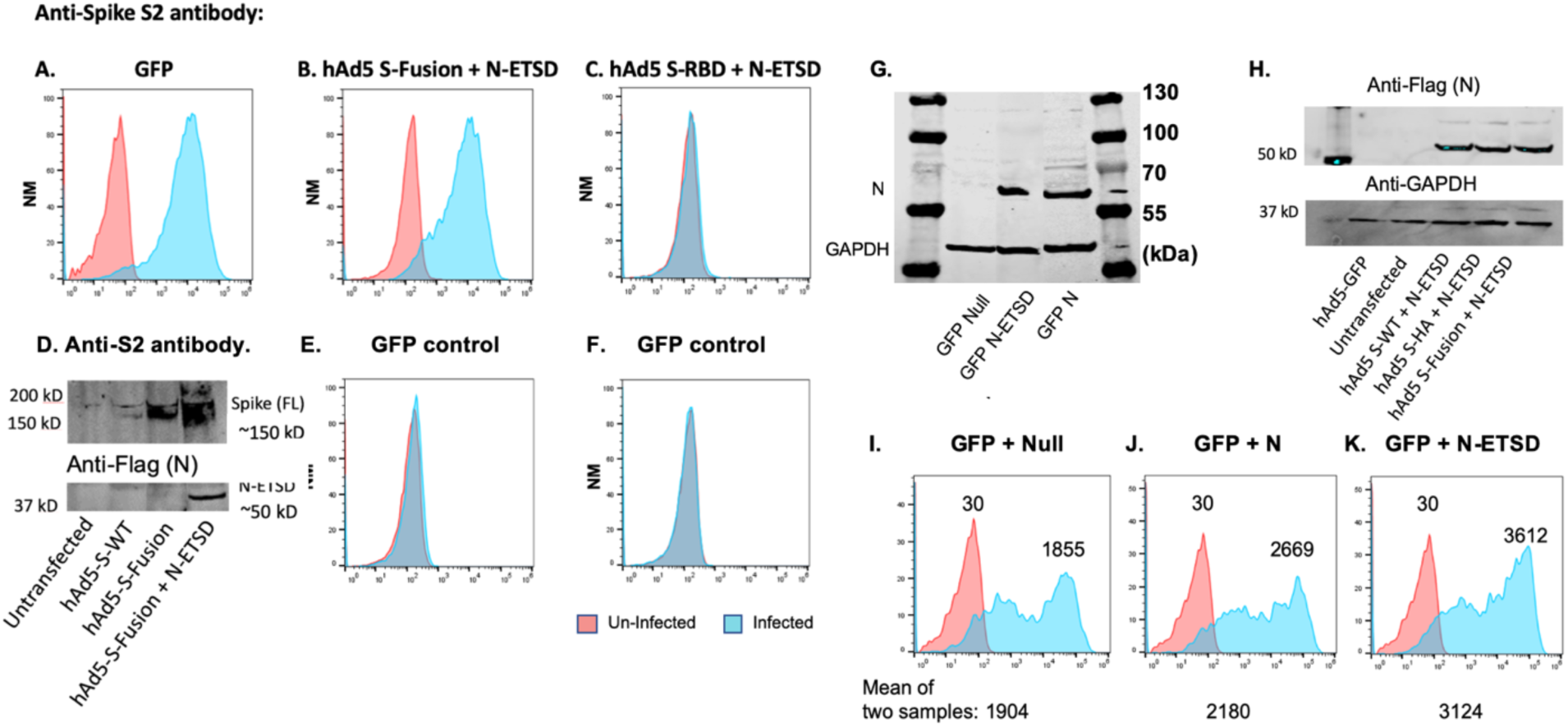
N-ETSD enhances protein co-expression. Anti-S2 antibody detection of untransfected (orange) and transfected (blue) HEK 293T cells for (A) GFP, (B) S-Fusion + N-ETSD and (C) negative control S RBD. (D) Anti-S2 probed immunoblot of cell lysates from untransfected, and hAd5 S-WT, S-Fusion, and S Fusion + N-ETSD transfected cells. (E, F) IgG controls. (G) Expression of N (∼65 kDa) in HEK 293T N-ETSD or N without ETSD transfected cells. The lower band is loading control GAPDH. (H) Co-expression of N-ETSD with other proteins (S-WT, S-HA, or S-Fusion) in cell lysate immunoblots probed with an anti-flag antibody recognizing the flag-tag on the N-ETSD construct. (I-K) GFP expression in HEK 293T cells transfected with GFP-expressing plasmids in combination with a null plasmid, N-WT, or N-ETSD; all at 250 ng for each (total of 500 ng). After 24 hours, GFP expression was evaluated by flow cytometry. As compared to co-transfection with a null plasmid, GFP (blue) was increased ∼1.4 fold with N-WT and almost 2-fold with N-ETSD (n = 2).

#### T-cell subpopulation phenotypes after enrichment

T-cell subpopulations of three previously infected SARS-CoV-2 patients were enriched using immunomagnetic bead selection (MojoSort BioLegend). The T cells were evaluated for co-receptor expression after enrichment using fluorescent conjugated antibodies to the co-receptors and cells acquired via flow cytometry (Fig. S5).

**Fig S5.**
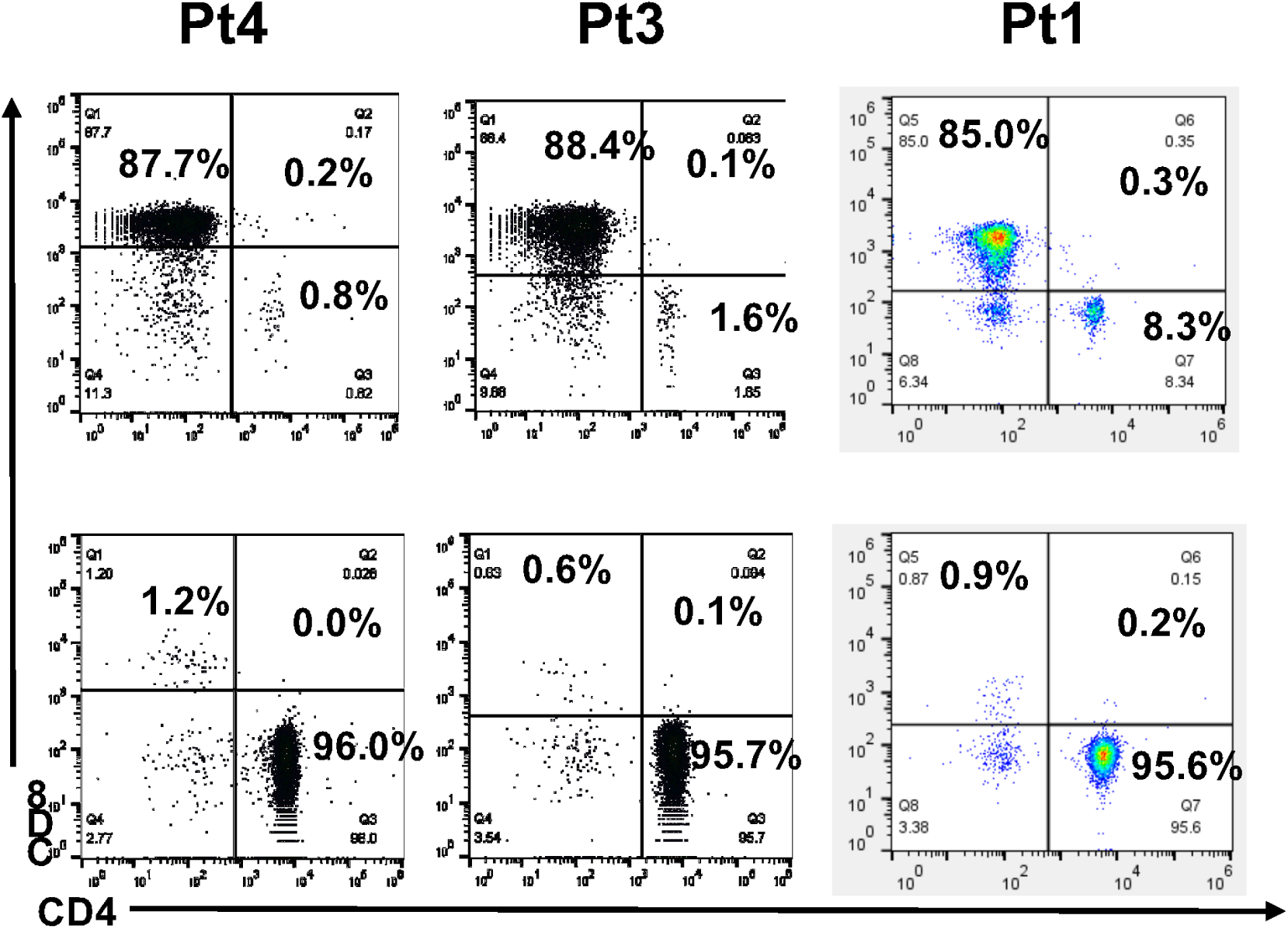
T-cell subpopulation phenotypes before and after enrichment. T cell subpopulations were enriched using immunomagnetic bead selection. The phenotype of the enriched T cells are shown above. Top row shows CD8+ enriched T cells and the bottom row displays CD4+ enriched T cells. Pt-previously SARS-CoV-2 infected patient blood donor.

## References

1 Sekine, T. et al. Robust T cell immunity in convalescent individuals with asymptomatic or mild COVID-19. bioRxiv, 2020.2006.2029.174888, doi:10.1101/2020.06.29.174888 (2020).

2 Le Bert, N. et al. SARS-CoV-2-specific T cell immunity in cases of COVID-19 and SARS, and uninfected controls. Nature, doi:10.1038/s41586-020-2550-z (2020).

3 Altmann, D. M. & Boyton, R. J. SARS-CoV-2 T cell immunity: Specificity, function, durability, and role in protection. Sci Immunol 5, doi:10.1126/sciimmunol.abd6160 (2020).

4 Matloubian, M., Concepcion, R. J. & Ahmed, R. CD4+ T cells are required to sustain CD8+ cytotoxic T-cell responses during chronic viral infection. Journal of virology 68, 8056–8063, doi:10.1128/jvi.68.12.8056-8063.1994 (1994).

5 Laidlaw, B. J., Craft, J. E. & Kaech, S. M. The multifaceted role of CD4(+) T cells in CD8(+) T cell memory. Nat Rev Immunol 16, 102–111, doi:10.1038/nri.2015.10 (2016).

6 Huang, C. et al. Clinical features of patients infected with 2019 novel coronavirus in Wuhan, China. The Lancet 395, 497–506, doi:10.1016/S0140-6736(20)30183-5 (2020).

7 Wang, C., Horby, P. W., Hayden, F. G. & Gao, G. F. A novel coronavirus outbreak of global health concern. The Lancet 395, 470–473, doi:10.1016/S0140-6736(20)30185-9 (2020).

8 Zhu, N. et al. A Novel Coronavirus from Patients with Pneumonia in China, 2019. New England Journal of Medicine 382, 727–733, doi:10.1056/NEJMoa2001017 (2020).

9 Chan, J. F.-W. et al. A familial cluster of pneumonia associated with the 2019 novel coronavirus indicating person-to-person transmission: a study of a family cluster. The Lancet 395, 514–523, doi:10.1016/S0140-6736(20)30154-9 (2020).

10 Hopkins, J. Cumulative Cases. Coronavirus Resource Center (2020).

11 Hopkins, J. Mortality Analyses. Coronavirus Resource Center (2020).

12 Rice, A. et al. A Next Generation Bivalent Human Ad5 COVID-19 Vaccine Delivering Both Spike and Nucleocapsid Antigens Elicits Th1 Dominant CD4+, CD8+ T-cell and Neutralizing Antibody Responses. bioRxiv, 2020.2007.2029.227595, doi:10.1101/2020.07.29.227595 (2020).

13 Zhu, F.-C. et al. Safety, tolerability, and immunogenicity of a recombinant adenovirus type-5 vectored COVID-19 vaccine: a dose-escalation, open-label, non-randomised, first-in-human trial. The Lancet, doi:10.1016/S0140-6736(20)31208-3.

14 van Doremalen, N. et al. ChAdOx1 nCoV-19 vaccination prevents SARS-CoV-2 pneumonia in rhesus macaques. bioRxiv, 2020.2005.2013.093195, doi:10.1101/2020.05.13.093195 (2020).

15 Amalfitano, A., Begy, C. R. & Chamberlain, J. S. Improved adenovirus packaging cell lines to support the growth of replication-defective gene-delivery vectors. Proceedings of the National Academy of Sciences of the United States of America 93, 3352–3356, doi:10.1073/pnas.93.8.3352 (1996).

16 Amalfitano, A. & Chamberlain, J. S. Isolation and characterization of packaging cell lines that coexpress the adenovirus E1, DNA polymerase, and preterminal proteins: implications for gene therapy. Gene Ther 4, 258–263, doi:10.1038/sj.gt.3300378 (1997).

17 Amalfitano, A. et al. Production and Characterization of Improved Adenovirus Vectors with the E1, E2b, and E3 Genes Deleted. Journal of virology 72, 926, doi:10.1128/JVI.72.2.926-933.1998 (1998).

18 Seregin, S. S. & Amalfitano, A. Overcoming pre-existing adenovirus immunity by genetic engineering of adenovirus-based vectors. Expert Opin Biol Ther 9, 1521–1531, doi:10.1517/14712590903307388 (2009).

19 Gatti-Mays, M. E. et al. A Phase I Trial Using a Multitargeted Recombinant Adenovirus 5 (CEA/MUC1/Brachyury)-Based Immunotherapy Vaccine Regimen in Patients with Advanced Cancer. Oncologist, doi:10.1634/theoncologist.2019-0608 (2019).

20 Gabitzsch, E. S. et al. Anti-tumor immunotherapy despite immunity to adenovirus using a novel adenoviral vector Ad5 [E1-, E2b-]-CEA. Cancer Immunology, Immunotherapy 59, 1131–1135, doi:10.1007/s00262-010-0847-8 (2010).

21 Lu, R. et al. Genomic characterisation and epidemiology of 2019 novel coronavirus: implications for virus origins and receptor binding. Lancet (London, England) 395, 565–574, doi:10.1016/S0140-6736(20)30251-8 (2020).

22 Wang, H. et al. The genetic sequence, origin, and diagnosis of SARS-CoV-2. Eur J Clin Microbiol Infect Dis, 1–7, doi:10.1007/s10096-020-03899-4 (2020).

23 Wu, A. et al. Genome Composition and Divergence of the Novel Coronavirus (2019-nCoV) Originating in China. Cell Host Microbe 27, 325–328, doi:10.1016/j.chom.2020.02.001 (2020).

24 Wrapp, D. et al. Cryo-EM structure of the 2019-nCoV spike in the prefusion conformation. Science 367, 1260–1263, doi:10.1126/science.abb2507 (2020).

25 Walls, A. C. et al. Structure, Function, and Antigenicity of the SARS-CoV-2 Spike Glycoprotein. Cell, doi:10.1016/j.cell.2020.02.058 (2020).

26 Kang, S. et al. Crystal structure of SARS-CoV-2 nucleocapsid protein RNA binding domain reveals potential unique drug targeting sites. Acta Pharmaceutica Sinica B, doi:https://doi.org/10.1016/j.apsb.2020.04.009 (2020).

27 Zeng, W. et al. Biochemical characterization of SARS-CoV-2 nucleocapsid protein. Biochemical and biophysical research communications, S0006-0291X(0020)30876-30877, doi:10.1016/j.bbrc.2020.04.136 (2020).

28 Srinivasan, S. et al. Structural Genomics of SARS-CoV-2 Indicates Evolutionary Conserved Functional Regions of Viral Proteins. Viruses 12, doi:10.3390/v12040360 (2020).

29 Burbelo, P. D. et al. Detection of Nucleocapsid Antibody to SARS-CoV-2 is More Sensitive than Antibody to Spike Protein in COVID-19 Patients. medRxiv: the preprint server for health sciences, 2020.2004.2020.20071423, doi:10.1101/2020.04.20.20071423 (2020).

30 Dutta, N. K., Mazumdar, K. & Gordy, J. T. The Nucleocapsid Protein of SARS-CoV-2: a Target for Vaccine Development. Journal of virology 94, e00647–00620, doi:10.1128/JVI.00647-20 (2020).

31 Hachim, A. et al. Beyond the Spike: identification of viral targets of the antibody response to SARS-CoV-2 in COVID-19 patients. medRxiv, 2020.2004.2030.20085670, doi:10.1101/2020.04.30.20085670 (2020).

32 Balint, J. P. et al. Extended evaluation of a phase 1/2 trial on dosing, safety, immunogenicity, and overall survival after immunizations with an advanced-generation Ad5 [E1-, E2b-]-CEA(6D) vaccine in late-stage colorectal cancer. Cancer immunology, immunotherapy: CII 64, 977–987, doi:10.1007/s00262-015-1706-4 (2015).

33 Korber, B. et al. Tracking Changes in SARS-CoV-2 Spike: Evidence that D614G Increases Infectivity of the COVID-19 Virus. Cell, doi:10.1016/j.cell.2020.06.043 (2020).

34 Dumonteil, E. & Herrera, C. Polymorphism and Selection Pressure of SARS-CoV-2 Vaccine and Diagnostic Antigens: Implications for Immune Evasion and Serologic Diagnostic Performance. Pathogens 9, doi:10.3390/pathogens9070584 (2020).

35 Peng, H. et al. Long-lived memory T lymphocyte responses against SARS coronavirus nucleocapsid protein in SARS-recovered patients. Virology 351, 466–475, doi:https://doi.org/10.1016/j.virol.2006.03.036 (2006).

36 Zhao, J. et al. Identification and Characterization of Dominant Helper T-Cell Epitopes in the Nucleocapsid Protein of Severe Acute Respiratory Syndrome Coronavirus. Journal of Virology 81, 6079–6088, doi:10.1128/jvi.02568-06 (2007).

37 Grifoni, A. et al. Targets of T Cell Responses to SARS-CoV-2 Coronavirus in Humans with COVID-19 Disease and Unexposed Individuals. Cell, doi:10.1016/j.cell.2020.05.015 (2020).

38 Shang, B. et al. Characterization and application of monoclonal antibodies against N protein of SARS-coronavirus. Biochemical and Biophysical Research Communications 336, 110–117, doi:https://doi.org/10.1016/j.bbrc.2005.08.032 (2005).

39 Azizi, A. et al. A combined nucleocapsid vaccine induces vigorous SARS-CD8+ T-cell immune responses. Genet Vaccines Ther 3, 7–7, doi:10.1186/1479-0556-3-7 (2005).

40 Narayanan, K., Chen, C.-J., Maeda, J. & Makino, S. Nucleocapsid-independent specific viral RNA packaging via viral envelope protein and viral RNA signal. Journal of virology 77, 2922–2927, doi:10.1128/jvi.77.5.2922-2927.2003 (2003).

41 McBride, R., van Zyl, M. & Fielding, B. C. The coronavirus nucleocapsid is a multifunctional protein. Viruses 6, 2991–3018, doi:10.3390/v6082991 (2014).

42 Nisreen, M. A. O. et al. Severe Acute Respiratory Syndrome Coronavirus 2−Specific Antibody Responses in Coronavirus Disease 2019 Patients. Emerging Infectious Disease journal 26, doi:10.3201/eid2607.200841 (2020).

43 Long, Q.-X. et al. Antibody responses to SARS-CoV-2 in patients with COVID-19. Nature Medicine, doi:10.1038/s41591-020-0897-1 (2020).

44 Tan, Y. J. et al. Profiles of antibody responses against severe acute respiratory syndrome coronavirus recombinant proteins and their potential use as diagnostic markers. Clin Diagn Lab Immunol 11, 362–371, doi:10.1128/cdli.11.2.362-371.2004 (2004).

45 Buchholz, U. J. et al. Contributions of the structural proteins of severe acute respiratory syndrome coronavirus to protective immunity. Proceedings of the National Academy of Sciences of the United States of America 101, 9804–9809, doi:10.1073/pnas.0403492101 (2004).

46 Montealegre, S. & van Endert, P. M. Endocytic Recycling of MHC Class I Molecules in Non-professional Antigen Presenting and Dendritic Cells. Front Immunol 9, doi:10.3389/fimmu.2018.03098 (2019).

47 Zhang, H., Penninger, J. M., Li, Y., Zhong, N. & Slutsky, A. S. Angiotensin-converting enzyme 2 (ACE2) as a SARS-CoV-2 receptor: molecular mechanisms and potential therapeutic target. Intensive Care Medicine, doi:10.1007/s00134-020-05985-9 (2020).

48 Lan, J. et al. Structure of the SARS-CoV-2 spike receptor-binding domain bound to the ACE2 receptor. Nature, doi:10.1038/s41586-020-2180-5 (2020).

49 Hoffmann, M. et al. SARS-CoV-2 Cell Entry Depends on ACE2 and TMPRSS2 and Is Blocked by a Clinically Proven Protease Inhibitor. Cell, doi:10.1016/j.cell.2020.02.052 (2020).

50 Tai, W. et al. Characterization of the receptor-binding domain (RBD) of 2019 novel coronavirus: implication for development of RBD protein as a viral attachment inhibitor and vaccine. Cell Mol Immunol, doi:10.1038/s41423-020-0400-4 (2020).

51 Suthar, M. S. et al. Rapid generation of neutralizing antibody responses in COVID-19 patients. Cell Reports Medicine, doi:10.1016/j.xcrm.2020.100040 (2020).

52 Brouwer, P. J. M. et al. Potent neutralizing antibodies from COVID-19 patients define multiple targets of vulnerability. Science, doi:10.1126/science.abc5902 (2020).

53 Robbiani, D. F. et al. Convergent antibody responses to SARS-CoV-2 in convalescent individuals. Nature 584, 437–442, doi:10.1038/s41586-020-2456-9 (2020).

54 Seow, J. et al. Longitudinal observation and decline of neutralizing antibody responses in the three months following SARS-CoV-2 infection in humans. Nature Microbiology, doi:10.1038/s41564-020-00813-8 (2020).

55 Ibarrondo, F. J. et al. Rapid Decay of Anti–SARS-CoV-2 Antibodies in Persons with Mild Covid-19. New England Journal of Medicine, doi:10.1056/NEJMc2025179 (2020).

56 Ng, O. W. et al. Memory T cell responses targeting the SARS coronavirus persist up to 11 years post-infection. Vaccine 34, 2008–2014, doi:10.1016/j.vaccine.2016.02.063 (2016).

57 Tang, F. et al. Lack of peripheral memory B cell responses in recovered patients with severe acute respiratory syndrome: a six-year follow-up study. Journal of immunology (Baltimore, Md.: 1950) 186, 7264–7268, doi:10.4049/jimmunol.0903490 (2011).

58 Channappanavar, R., Fett, C., Zhao, J., Meyerholz, D. K. & Perlman, S. Virus-specific memory CD8 T cells provide substantial protection from lethal severe acute respiratory syndrome coronavirus infection. Journal of virology 88, 11034–11044, doi:10.1128/jvi.01505-14 (2014).

59 Zhou, R. et al. Acute SARS-CoV-2 Infection Impairs Dendritic Cell and T Cell Responses. Immunity 53, 864-877.e865, doi:10.1016/j.immuni.2020.07.026 (2020).

60 Ferretti, A. P. et al. Unbiased screens show CD8+ T cells of COVID-19 patients recognize shared epitopes in SARS-CoV-2, most of which are not located in the Spike protein. Immunity, doi:https://doi.org/10.1016/j.immuni.2020.10.006 (2020).

61 Kabeya, Y. et al. LC3, a mammalian homologue of yeast Apg8p, is localized in autophagosome membranes after processing. Embo j 19, 5720–5728, doi:10.1093/emboj/19.21.5720 (2000).

62 Schmid, D., Pypaert, M. & Münz, C. Antigen-loading compartments for major histocompatibility complex class II molecules continuously receive input from autophagosomes. Immunity 26, 79–92, doi:10.1016/j.immuni.2006.10.018 (2007).

63 Patterson, N. L. & Mintern, J. D. Intersection of autophagy with pathways of antigen presentation. Protein & Cell 3, 911–920, doi:10.1007/s13238-012-2097-3 (2012).

64 Crotzer, V. L. & Blum, J. S. Autophagy and Its Role in MHC-Mediated Antigen Presentation. The Journal of Immunology 182, 3335, doi:10.4049/jimmunol.0803458 (2009).

65 You, L. et al. The crosstalk between autophagic and endo-/exosomal pathways in antigen processing for MHC presentation in anticancer T cell immune responses. Journal of Hematology & Oncology 10, 165, doi:10.1186/s13045-017-0534-8 (2017).

66 Tan, C. W. et al. A SARS-CoV-2 surrogate virus neutralization test based on antibody-mediated blockage of ACE2-spike protein-protein interaction. Nat Biotechnol, doi:10.1038/s41587-020-0631-z (2020).

67 Almazán, F., Galán, C. & Enjuanes, L. The Nucleoprotein Is Required for Efficient Coronavirus Genome Replication. Journal of Virology 78, 12683–12688, doi:10.1128/jvi.78.22.12683-12688.2004 (2004).

68 Bennett, S. R. M. et al. Help for cytotoxic-T-cell responses is mediated by CD40 signalling. Nature 393, 478–480 (1998).

69 Schoenberger, S. P., Toes, R. E. M., van der Voort, E. I. H., Offringa, R. & Melief, C. J. M. T-cell help for cytotoxic T lymphocytes is mediated by CD40-CD40L interactions. Nature 393, 480–483 (1998).

70 Niazi, K. R. et al. Activation of human CD4+T cells by targeting MHC class II epitopes to endosomal compartments using human CD1 tail sequences. Immunology 122, 522–531, doi:10.1111/j.1365-2567.2007.02666.x (2007).

71 Gupta, V. et al. SARS coronavirus nucleocapsid immunodominant T-cell epitope cluster is common to both exogenous recombinant and endogenous DNA-encoded immunogens. Virology 347, 127–139, doi:10.1016/j.virol.2005.11.042 (2006).

72 Lőrincz, P. & Juhász, G. Autophagosome-Lysosome Fusion. Journal of Molecular Biology 432, 2462–2482, doi:https://doi.org/10.1016/j.jmb.2019.10.028 (2020).

73 Yang, K. et al. Immune responses to T-cell epitopes of SARS CoV-N protein are enhanced by N immunization with a chimera of lysosome-associated membrane protein. Gene Ther 16, 1353–1362, doi:10.1038/gt.2009.92 (2009).

74 Soresina, A. et al. Two X-linked agammaglobulinemia patients develop pneumonia as COVID-19 manifestation but recover. Pediatric Allergy and Immunology 31, 565–569, doi:10.1111/pai.13263 (2020).

75 Moderbacher, C. R. et al. Antigen-specific adaptive immunity to SARS-CoV-2 in acute COVID-19 and associations with age and disease severity. Cell, doi:10.1016/j.cell.2020.09.038 (2020).

76 Kroemer, M. et al. COVID-19 patients display distinct SARS-CoV-2 specific T-cell responses according to disease severity. The Journal of infection, 4816–4816, doi:10.1016/j.jinf.2020.08.036 (2020).

77 Chen, Z. & John Wherry, E. T cell responses in patients with COVID-19. Nat Rev Immunol, 1–8, doi:10.1038/s41577-020-0402-6 (2020).

78 Weiskopf, D. et al. Phenotype and kinetics of SARS-CoV-2–specific T cells in COVID-19 patients with acute respiratory distress syndrome. Science Immunology 5, eabd2071, doi:10.1126/sciimmunol.abd2071 (2020).

79 Sin, J.-I. et al. IL-12 Gene as a DNA Vaccine Adjuvant in a Herpes Mouse Model: IL-12 Enhances Th1-Type CD4<sup>+</sup> T Cell-Mediated Protective Immunity Against Herpes Simplex Virus-2 Challenge. The Journal of Immunology 162, 2912–2921 (1999).

80 Channappanavar, R., Fett, C., Zhao, J., Meyerholz, D. K. & Perlman, S. Virus-Specific Memory CD8 T Cells Provide Substantial Protection from Lethal Severe Acute Respiratory Syndrome Coronavirus Infection. Journal of Virology 88, 11034–11044, doi:10.1128/jvi.01505-14 (2014).

81 Kennedy, R. C., Shearer, M. H., Watts, A. M. & Bright, R. K. CD4+ T Lymphocytes Play a Critical Role in Antibody Production and Tumor Immunity against Simian Virus 40 Large Tumor Antigen. Cancer research 63, 1040 (2003).

82 Swain, S. L., McKinstry, K. K. & Strutt, T. M. Expanding roles for CD4+ T cells in immunity to viruses. Nat Rev Immunol 12, 136–148, doi:10.1038/nri3152 (2012).

83 Braun, J. et al. SARS-CoV-2-reactive T cells in healthy donors and patients with COVID-19. Nature, doi:10.1038/s41586-020-2598-9 (2020).

84 Schmidt, M. E. & Varga, S. M. The CD8 T Cell Response to Respiratory Virus Infections. Front Immunol 9, doi:10.3389/fimmu.2018.00678 (2018).

85 Peng, Y. et al. Broad and strong memory CD4+ and CD8+ T cells induced by SARS-CoV-2 in UK convalescent COVID-19 patients. bioRxiv https://doi.org/10.1101/2020.06.05.134551, 2020.2006.2005.134551, doi:10.1101/2020.06.05.134551 (2020).

86 Kasinrerk, W., Baumruker, T., Majdic, O., Knapp, W. & Stockinger, H. CD1 molecule expression on human monocytes induced by granulocyte-macrophage colony-stimulating factor. The Journal of Immunology 150, 579–584 (1993).

